# Healthcare visits of the population 50+ in Europe from 2004 to 2022

**DOI:** 10.1101/2024.07.10.24310226

**Authors:** Van Kinh Nguyen, Anna Reuter, Mirna Abd El Aziz, Till Bärnighausen

## Abstract

**Background:** The COVID-19 pandemic brought major reductions in healthcare visits in Europe. The long-run consequences for healthcare systems depend on the paths to post-pandemic levels, especially among the groups in highest need of sustained care.

**Methods:** We used individual longitudinal data from 27 European countries from the Survey of Health, Ageing and Retirement in Europe, divided in three phases: pre-pandemic (2004-2019), pandemic (2021), and post-pandemic (2022). We analysed the number of healthcare visits by gender over time and their correlation with age and chronic conditions using a Bayesian spatio-temporal model.

**Results:** Before COVID-19, the nominal rate of healthcare visits varied among countries from two to six visits per year and increased over time. The COVID-19 pandemic severely affected the number of healthcare visits, with an estimated reduction from 65% to 95% across countries. During COVID-19, older individuals and those with chronic conditions had a more than proportional reduction in the number of healthcare visits. After COVID-19, the pattern of healthcare visits mostly returned to pre-pandemic levels, with an over-shooting for people with cancer and a lag for people with cardiovascular diseases.

**Conclusion:** The quick recovery in the number of healthcare visits for most European countries indicate that access to care stabilised again. Yet, the different trends for vulnerable groups show that these groups require timely action to prevent long-term consequences of missed care.

## Introduction

The COVID-19 pandemic brought severe disruptions to the healthcare systems worldwide. On the one hand, even relatively resource-rich healthcare systems as in Europe were overwhelmed by the number of intensive care patients and the high infection risk. On the other hand, individuals stayed absent due to fear of infection, mobility restrictions, or rejection by the healthcare providers. In some cases, this might have relieved healthcare systems from overutilization in terms of unnecessary consultations and treatments [1], particularly in systems with a high coverage of health insurance, as in many European countries. Yet, if diagnoses were delayed or chronic care interrupted, the long-run consequences could put an immense burden on the healthcare systems in the near future. The massive drop in healthcare utilization, especially for outpatient care, during the early phase of the pandemic has been widely documented [2, 3, 4, 5]. While some studies indicate larger reductions among patients in better health [2], others point towards a delay in diagnoses for cardiovascular diseases, diabetes, cancer, and common mental health problems [6, 7, 8]. For delayed melanoma diagnoses alone, it is estimated that $7.65 billion additional costs incurred in Europe [9].

There is few evidence on the trajectory of outpatient care after the initial phase of the pandemic, and most covering periods till 2021 only. In the UK, care for common mental health problems seem to have recovered to pre-pandemic levels by 2021 [10], as did visits to general practitioners and specialists in Germany [11]. In Poland, the number of healthcare users of primary care, specialist care, and preventive programs recovered in 2021, but not the volume of care [12]. In Norway, specialists bookings recovered, but primary care bookings stayed below pre-pandemic levels in 2021 [13]. Similarly, wellness visits and preventive screenings in 2022 remained below pre-pandemic levels in the US [14]. The impact on disparities in access to health is less clear, but among the older population in Europe, those with poorer health were more likely to report missed healthcare visits during the pandemic [15].

We aimed to investigate the trajectories of healthcare visits in Europe before, during and after the pandemic, and shed light on the disparities among the population most in need of sustained healthcare. We focused on the population aged 50 and older, which is particularly in need of regular healthcare based on the high prevalence of chronic conditions and represented in the Survey of Health, Ageing and Retirement in Europe (SHARE). The longitudinal structure allows us to estimate the changes in the number of healthcare visits (NHV) during the pandemic (year 2021) and after the pandemic (years 2021/2022) in comparison to the pre-pandemic period (years 2004-2019), and to analyze the disparities by age, gender, and diagnosed health conditions.

## Materials and Methods

### Data

Data from 27 European countries were collated from the Survey of Health, Ageing and Retirement in Europe (SHARE), waves 1, 2, 4, 5, 6, 7, 8, 9 [16, 17, 18, 19, 20, 21, 22, 23] and the second wave of the COVID-19 survey [24]. The SHARE panel includes health and socioeconomic data of respondents aged 50 or older and their partners. The first wave was conducted in 2004, spanning 11 European countries and Israel. The following waves were conducted approximately every two years, and over time further 16 European countries joined SHARE. In March 2020, data collection for wave 8 had to be stopped due to the outbreak of the COVID-19 pandemic. In summer 2020 and summer 2021, two special COVID-19 surveys were conducted over the phone. From October 2021 till December 2022, wave 9 was conducted (with 80% of interviews in 2022), returning back to the standard questionnaire and mode. In the following, we refer to the COVID-19 survey as data from the “pandemic” period, and the wave 9 data as data from the “post-pandemic” period.

The primary outcome was the number of healthcare visits an individual reported in the period prior to the interview (NHV). Healthcare visits were considered as seeing or talking to a medical doctor or qualified nurse, excluding visits to dentists and hospital stays, but including emergency or outpatient clinic visits. In the COVID-19 survey, healthcare visits were recorded as going to a doctor’s office or a medical facility other than a hospital. The recall period was 12 months for waves 1-9, and since the last interview/past 12 months for the second COVID-19 survey. For this wave, we used the exact date of interview to calculate the person-month contributed by each individual. For the first COVID-19 survey, the recall period was defined as ’since the COVID-19 outbreak’, which might vary based on the respondent’s perception on the beginning of the outbreak, and was hence excluded from the analysis. Details can be found in the Appendix S1.

We excluded respondents who were staying at a nursing facility at the time of interview, those younger than 50 years at the time of the interview (these are partners of the main respondents), as they were not considered as target sample, and respondents aged 100 or older to preclude selection effects at high ages. To retain the maximum sample size with an observed number of healthcare visits, we matched individuals across the survey waves and imputed missing values for the covariates (age and gender) by records in the other waves. Existing health conditions were captured as self-reported diagnoses according to a pre-specified list. Health conditions that were collected only in the most recent waves were excluded from the analysis. Ireland participated in only one survey and was not included in this analysis. Overall, data from 147,116 individuals, resulting in 505,607 observations, were included in the analyses.

In addition, we used the following additional country level data to analyse differences across countries: GDP per capita in 2019 [25], government health expenditure as % of GDP in 2019 [26], physician density in 2013 (latest date available) [27], population density in 2019 [27], cumulative deaths by June 2021 [28], and the average stringency index till June 2021 by the Oxford Government Response Tracker [28].

### Model and implementation

We implemented a combined likelihoods approach to estimate the covariate effect without the influence of epidemic and post-epidemic data, as well as providing a concrete reference for the estimation of the epidemic effects. First, the number of healthcare visits during a time period was modelled as a rate per person-month. This allows different periods of recall to be modelled and projected. We constructed a Bayesian hierarchical model for the number of healthcare visit *y_ijt_* in the corresponding observation period *E_ijt_* months for an individual *i* in country *j* at interview time *t* as follows:

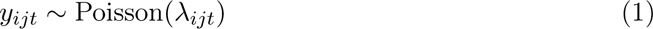

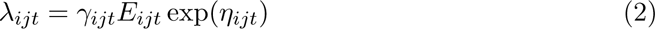

In particular, we modelled the number of healthcare visits as a Poisson count that was extended to allow a larger variation of the NHV via the over-dispersion parameter (*γ*) with a unit prior mean, and separately for men and women. For the pre-epidemic data period, the linear predictor is modelled as

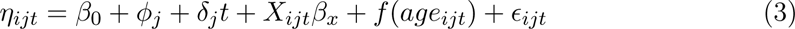

where we assumed a country-specific difference, *ϕ_j_* ∼ *N* (0*, σ*^2^), that varied around the global mean *β*_0_. The mean NHV varied according to a set of health conditions with a shared fixed-effect among the countries, *X_ijt_β_x_*. A shared effect of the continuous age variable was modelled with a second-order random walk to allow for potential structured nonlinear effects, denoted by *f* (*age_ijt_*). Trends over time, represented by the year of interview, were modelled with a random slope term *δ_j_*, which allows for differences in the trends among the countries. We assumed that the trend among the countries might be correlated and modelled the slopes with the spatially correlated model BYM [29].

Individual identification across the waves was modelled with a normally distributed term 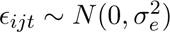 to take into account within-individual variability. We used only the data before the COVID-19 pandemic to inform the model’s parameters at this step. We tested for beta convergence by regressing the yearly growth rate of the NHV on the initial NHV. For the epidemic period, the estimated linear predictor from the pre-epidemic model was carried over to provide the reference point for the case without an epidemic. The data of this period provides estimates of the changes compared to what would be predicted given the same individual characteristics. The linear predictor for this period of time *u* read as

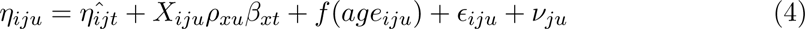

where 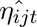 denotes the expected level of NHV without epidemic, *ρ_xu_* models the relative change of a comorbidity effect compared to pre-epidemic, a separate age pattern during the epidemic was modeled with the RW2 term *f* (*age_iju_*), and *ν_ju_* denotes the country-specific change in the mean of the NHV during the epidemic. Similarly, for the post-epidemic period, we assumed that there might exist a carry-over effect of the pandemic and replaced the coefficients in Equation 4 with those representing the estimates from wave 9.

All equations were jointly estimated, using an augmented data approach and three likelihoods for the three data periods, in which the parameters were copied over to the epidemic and post-epidemic periods, including their uncertainty in the estimates [30].

There were three parts of the data, each of which yields a slightly different data likelihood. For the survey waves 1, 2, 4, and 5, SHARE recorded the NHV truncated at the value 98; during waves 6 − 9, SHARE recorded the NHV without truncation; during the second COVID-19 survey, SHARE recorded the NHV as a binary response, which we interpreted as a Poisson variable truncated at one. The three parts can be modelled as a censored Poisson at 98, a Poisson, and a censored Poisson at one, respectively. The likelihood reads as

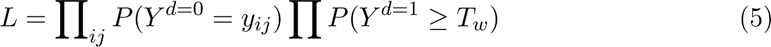

where *d* = 0, 1 is an indicator whether the observation was censored; *T_w_* = {1, 98} denotes two cases of censored data described above. The model parameters were estimated with the R package INLA [31]. We computed posterior medians and 95% credible intervals of the outcomes of interests, including the annual NHV, the differential of the time trend, and the effect of COVID-19, using posterior simulations of 1000 parameter samples. The models were compared using the Watanabe Information Criteria (WAIC). Computations were done using the High Performance Computer Cluster at the Center for Scientific Computing (CSC) of the Goethe University Frankfurt with the single model runtime 2̃6h on 40-cores 128Gb RAM AlmaLinux.

## Results

### Healthcare visits before the pandemic

Fig. 1 shows the expected annual number of healthcare visits by country and sex estimated for the year 2019, controlling for age, comorbidities, and trends over time. The nominal rate of visits, i.e., for an individual without comorbidites and of average age 75 years, varied from 1.5 visits to 5.6 visits per year, and was higher for women than for men. By 2019, the average NHV across countries was 3.5 for women and 3.0 for men. The highest rate of visits in 2019 was observed in the centre of Europe, with 5.6 nominal visit for women and 4.6 nominal visits for men in Luxembourg, followed by Belgium (women: 5.1, men: 4.3), Italy (women: 4.6, men: 4.0), Austria (women: 4.5,men: 4.0), and Germany (women: 4.5, men: 4.0). The lowest rate of visits was observed in Northern Europe and small islands, with 2.2 nominal visits for women and 2.0 nominal visits for men in Finland, followed by Cyprus (women: 2.6, men: 2.0), and Malta (women: 2.5, men: 2.2).

**Figure 1:**
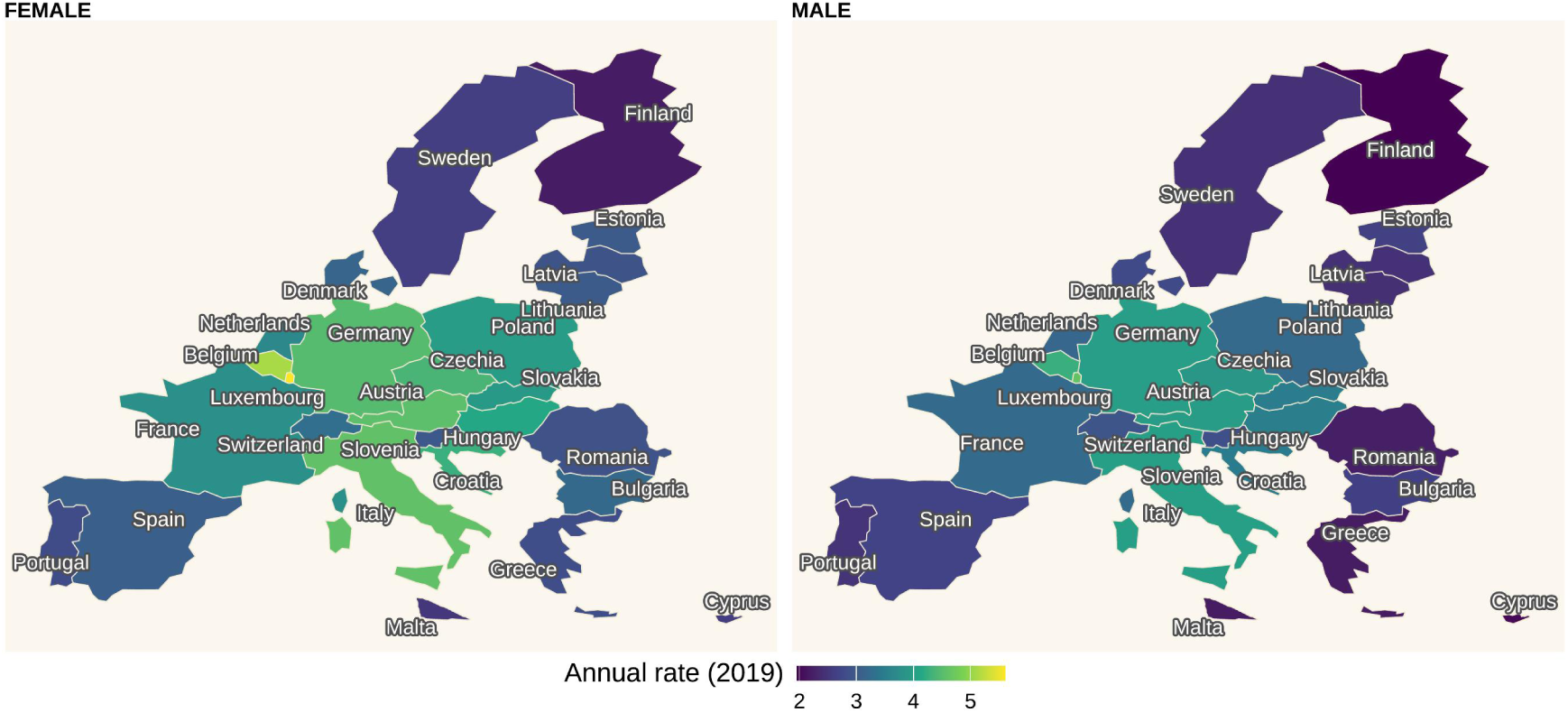
Posterior estimate of the rate of healthcare visits annualised at the year 2019. The estimates are conditioned on the reference group in the regression model: no comorbidity and an average age (approx. 75)

Across Europe, there was a large variation in trends in NHV over time, with a slight increase overall (Fig. S1). There was a small convergence, with countries with initially higher rates experiencing a smaller increase in the NHV than countries with initially lower rates (beta convergence men: -2.3%, p-value 0.081, beta convergence women: -2.4%, p-value 0.023). Within most countries, trends were relatively similar for women and men, while for some countries, such as Italy, Spain, France, and Czechia the gender gap in NHV narrowed over time.

### Pandemic shock and recovery

Fig. 2 shows the estimated relative reduction of the healthcare visits rate during and after the COVID-19 pandemic by country. The estimates were derived from the difference between the expected rate if the pandemic had not occurred based on the pre-epidemic data. Individuals in all countries experienced a large drop in the rate of healthcare visits (Fig. 2); the least affected country was Germany with an estimated reduction of 55% for men and 58% for women. The most affected countries were Italy, Croatia, Estonia, and Hungary, where the healthcare visits came nearly to a halt with an estimated reduction of up to 95% in the NHV. In 2022, healthcare visits returned back to the pre-pandemic levels for most of the countries, with a NHV between 90% and 110% compared to prepandemic levels. The rate still remained low in Slovakia (men: 64%, women: 71%), Luxembourg (men: 81%, women: 85%), and Poland (men: 85%, women: 80%), and was relatively high compared to pre-pandemic levels in Cyprus (men: 136%, women: 114%).

**Figure 2:**
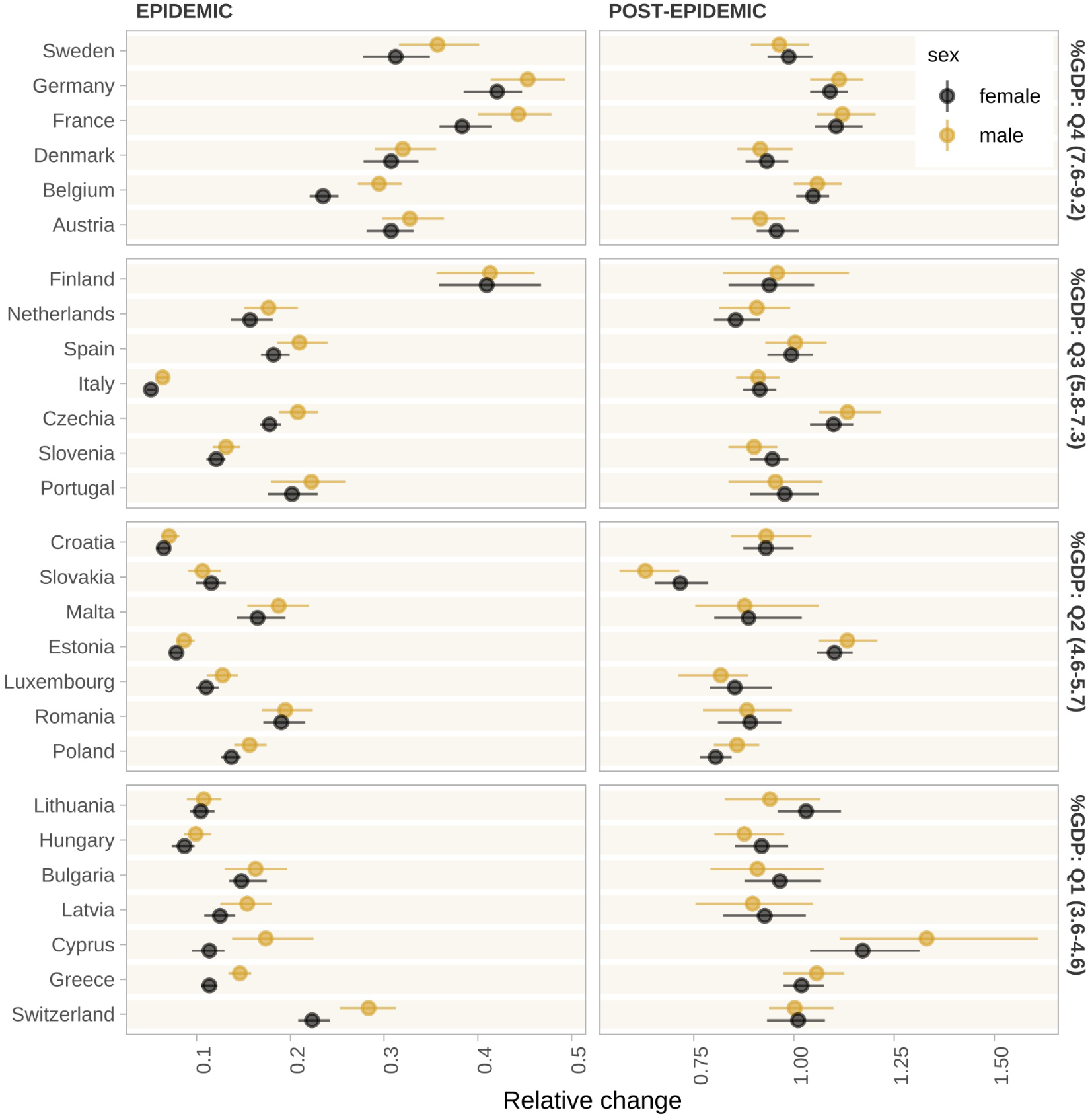
Posterior estimate of the percentage reduction of the healthcare visits rate compared to what is expected by the past trend, the point and number are the posterior median, the lines are the 95% uncertainty interval. The countries are grouped by percent of domestic general government health expenditure of GDP for visualization purposes.

We grouped the countries according to the government health expenditure as % of GDP in 2019. Countries with a very high health expenditure tended to have a smaller decline of healthcare visits during COVID-19, compared to those with a lower health expenditure. Overall, a 1 percentage point increase in government health expenditure as % of GDP in 2019 was correlated with a 4.7% higher NHV during the pandemic compared to pre-pandemic levels (i.e., a smaller reduction) for men and a 4.5% higher NHV during the pandemic compared to pre-pandemic levels for women. Neither the pandemic drop nor the post-pandemic recovery correlated with the severity of the pandemic (cumulative deaths or average Oxford Government Response stringency index by the time of the second COVID-19 survey), the population density in 2019, or the physician density in 2013. If Luxembourg, as an extreme outlier, was omitted, there was a negative correlation between the pandemic decline and GDP p.c. in 2019.

### Healthcare visits across age and comorbidities

Fig. 3 shows that most chronic conditions were associated with a higher rate of healthcare visits. The largest difference in the rate of healthcare visits was among people with cancer compared to people without cancer. Having chronic lung diseases, diabetes, a history of stroke, or heart attack was associated with an increase in the visit rate by approximately 20% compared to those without the disease. For most health conditions, the correlation was larger for men than for women. For stroke, stomach and related ulcers and high cholesterol, the correlation was similarly high for both sexes. During the epidemic period, the NHV decreased relatively more for most of the chronic conditions, but remained slightly higher than for respondents without a condition. After the pandemic, the correlation with NHV remained significantly lower for men with a prior heart attack (relative risk post-epidemic 1.30, 95% CI [1.29 − 1.31], pre-epidemic 1.48 [1.46 − 1.50]), hypertension (post: 1.29 [1.28 − 1.30], pre: 1.34 [1.33 − 1.36]), diabetes (post: 1.31 [1.29 − 1.32], pre: 1.43 [1.41 − 1.46]), and chronic lung disease (post: 1.29 [1.27 − 1.31], pre: 1.42 [1.39 − 1.44]), but was significantly higher for men with a cancer diagnosis (post: 1.90 [1.87 − 1.94], pre: 1.78 [1.75 − 1.81]) and stomach, duodenal, or peptic ulcer (post: 1.28 [1.24 − 1.32], pre: 1.19 [1.16 − 1.21]). For women, the picture was similar, except for ulcers: The correlation remained significantly lower for women with a prior heart attack (post: 1.30 [1.28 − 1.31], pre: 1.34 [1.32 − 1.36]), hypertension (post: 1.24 [1.23 − 1.25], pre: 1.29 [1.28 − 1.31], stroke (post: 1.21 [1.19 − 1.22], pre: 1.30 [1.26 − 1.32]), diabetes (post: 1.23 [1.22 − 1.25], pre: 1.34 [1.32 − 1.36]), and chronic lung disease (post: 1.27 [1.25 − 1.29], pre: 1.32 [1.30 − 1.34]), but significantly higher for cancer (post: 1.70 [1.67 − 1.72], pre: 1.62 [1.59 − 1.64]).

**Figure 3:**
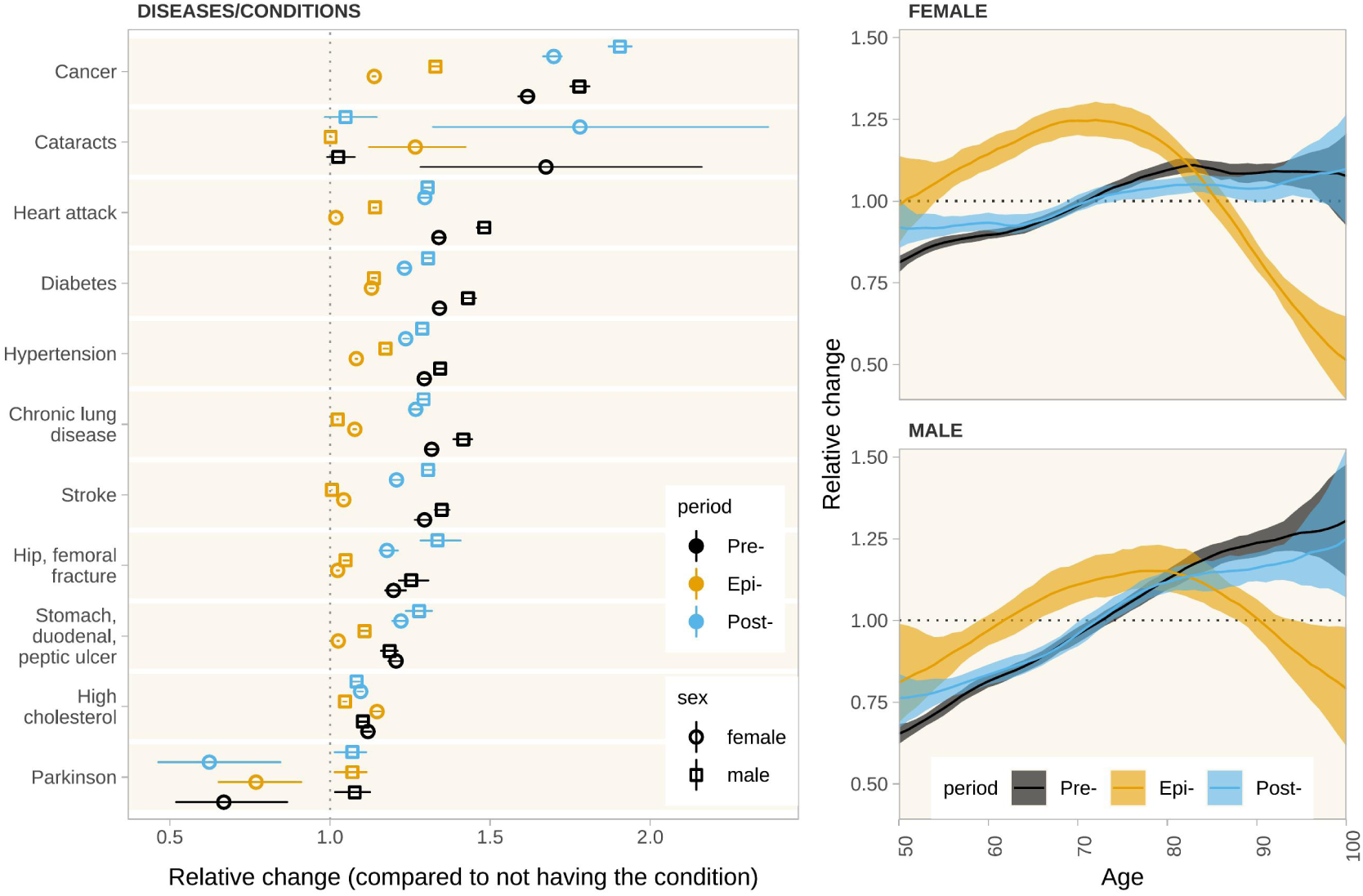
Posterior estimate of the relative rate of healthcare visits by sex. The point/line and line range/shaded colours denote the posterior median and 95% uncertainty interval, respectively. The diseases and health conditions were compared against not being diagnosed with the respective condition. Age was modelled as nonlinear terms for each of the periods (pre, epi, post) separately.

During the pre-epidemic period, age was positively linearly correlated with the number of visits, with a steeper trend for men compared to women. This pattern, however, changed during the epidemic into an inverse u-shaped relationship: For respondents aged 50-75, age was positively correlated with the NHV; but for respondents at higher ages, there was a steep negative correlation. After the pandemic, the positive linear correlation returned for both sexes, although with slightly flatter slopes, particularly for women.

## Discussion

Evidence on the trajectories of healthcare visits after the pandemic is scarce for most European countries, especially for disaggregated data. We made use of a harmonized, longitudinal dataset of 147,116 individuals aged 50 and older in Europe, and found a large decline in the number of healthcare visits during the COVID-19 pandemic. While the extent of the decline differed across countries, age groups, and by prior diagnoses, the major share of the decline was reduced quickly after 2021. For people with prior diagnoses, relative disparities in the number of healthcare visits persisted, pointing towards a backlog in the healthcare systems. The pre-pandemic, positive age gradient in healthcare visits is in line with previous findings on the population aged 50 and older in Europe [32, 33]. The correlation might be driven by the accumulation of health deficits as people age [34], national screening guidelines, or might proxy time to death [35]. We found that during the pandemic, individuals above 75 reduced their healthcare visits more than proportionally, such that the age gradient reversed. At the same time, another study found that the incidence of missed healthcare visits due to fear of COVID-19, postponed, or denied appointments, decreased with age [15]. This discrepancy might be explained either by the definition of “missed healthcare”, which did not capture other reasons such as mobility restrictions or (perceived) recommendations to stay at home, especially given the higher risk of severe infections due to higher age, or by the respondents’ perceptions, e.g. to which extent “fear” of infection drove the decision to not make an appointment. While the age gradient returned to a linear form after the pandemic, the correlation was flatter than before the pandemic, such that individuals at high ages faced a slightly lower relative NHV. The age gradient for the younger respondents remained quite similar during the pandemic, but the NHV were relatively higher after the pandemic, indicating a small catch-up effect for the population aged 50-60. This might point towards a backlog in general health checks and screenings, as this age group is at the brink (or just above) several screening guidelines [36, 37]. Indeed, findings from England show that blood pressure screening rates decreased for middle-aged individuals during the pandemic [38]. Previous studies on SHARE data primarily analysed the number of diagnoses of condition, or deficit indices containing diagnosed conditions, finding a positive correlation between multimorbidity and healthcare visits [39, 33, 40]. We showed that the difference in healthcare visits among diagnosed and undiagnosed respondents was particularly high for cancer, and comparatively low for stomach and related ulcers and high cholesterol. During COVID-19, these associations decreased, resulting in a relatively larger gap in care for people with chronic conditions. There was no clear pattern according to the type of condition, e.g., whether patients were at high risk of a severe infection. The NHV increased more than proportionally for individuals with cancer after COVID-19, indicating that their conditions might have become more severe. This is similar to findings from the UK, where median attendance volume decreased for cancer patients during the lockdowns, followed by large increases in consultation rate afterwards [41]. At the same time, the NHV increased less than proportionally for (mostly) less acute conditions such as diabetes, hypertension, or a former heart attack or stroke, such that these individuals still experienced a gap in care compared to the time before the pandemic. Without further attention to ensure a full retention in care, these conditions are likely to worsen over time and induce higher healthcare costs in the medium run. Most European countries, as many countries worldwide, reacted to the onset of the pandemic with sudden and strict lockdown policies [28]. Yet, there was a large heterogeneity in the timing of large outbreaks, pandemic preparedness, and pandemic management, and finally in the excess mortality during the pandemic [42]. Health expenditure and GDP per capita, but not the average stringency index or population density were correlated with excess mortality [42], which mirrors our findings on NHV. Despite these heterogeneities, most countries returned to levels of healthcare visits similar to those before the pandemic.

Our study comes with several limitations. The number of healthcare visits measures healthcare access and utilization, but not necessarily whether utilization is adequate. As the number of objectively needed healthcare visits is individual and cannot be observed, we cannot distinguish between underutilization, adequate utilization, and overutilization. Hence, a higher rate of healthcare visits does not necessarily reflect better healthcare quality. Still, disparities in the rate of healthcare visits across groups and over time hint at potential deficits for one of the groups, and thus indicate the need for further investigation and potential policy intervention. Similarly, as healthcare visits are self-reported, we cannot rule out potential reporting biases common to survey data. Another limitation is the changed measurement of healthcare utilization during COVID-19. First, only the extensive margin, i.e., whether a doctor was visited, was assessed. Second, the phrasing of the question differed, as respondents were asked whether they went to a facility other than a hospital, compared to whether they have seen or talked to a doctor or nurse in the previous waves. We tried to incorporate these differences by modelling different Poisson distributions, but cannot rule out that this led to systematic differences in measurement which cannot be captured by the model. However, given that wave 9 returned to the previous questionnaire, we are confident that the reported post-COVID changes accurately reflect the difference to the pre-COVID period.

Overall, our findings suggest that while access to care largely returned to pre-pandemic levels for the population age 50+ in Europe and across countries, people at higher ages and with chronic conditions require more attention from research and policy making. Both groups were disproportionately affected in their access to care, despite being the groups most in need of regular healthcare visits. In the aftermath of this pandemic, their retention-to-care needs to be ensured, and more monitoring might be required to detect health consequences of the disruption in care early on. For future pandemics, strategies are needed to ensure that disruptions in care are particularly monitored and buffered for those vulnerable groups.

## Data Availability

Access to the data can be granted by the SHARE project (http://share-eric.eu/).

https://share-eric.eu/data/

## Declarations

### Author contributions

AR conceptualized the study, VKN developed the methods and analyzed the data. VKN and AR wrote the first draft of the manuscript. All authors reviewed and edited the final draft.

## Acknowledgments

We acknowledge Jeff Imai-Eaton for suggesting the use of truncated Poisson to account for differences between normal waves and COVID waves.

This paper uses data from SHARE Waves 1, 2, 4, 5, 6, 7, 8 and 9 as well as SHARE Corona Survey 2 [16, 17, 18, 19, 20, 21, 22, 23, 24], see Börsch-Supan et al. (2013) and Scherpenzeel et al. (2020) for methodological details [43, 44]. The SHARE data collection has been funded by the European Commission, DG RTD through FP5 (QLK6-CT-2001-00360), FP6 (SHARE-I3: RII-CT-2006-062193, COMPARE: CIT5-CT-2005-028857, SHARELIFE: CIT4-CT-2006-028812), FP7 (SHAREPREP: GA N°211909, SHARE-LEAP: GA N°227822, SHARE M4: GA N°261982, DASISH: GA N°283646) and Horizon 2020 (SHARE-DEV3: GA N°676536, SHARE-COHESION: GA N°870628, SERISS: GA N°654221, SSHOC: GA N°823782) and by DG Employment, Social Affairs & Inclusion through VS 2015/0195, VS 2016/0135, VS 2018/0285, VS 2019/0332, and VS 2020/0313. Additional funding from the German Ministry of Education and Research, the Max Planck Society for the Advancement of Science, the U.S. National Institute on Aging (U01_AG09740-13S2, P01_AG005842, P01_AG08291, P30_AG12815, R21_AG025169, Y1-AG-4553-01, IAG_BSR06-11, OGHA_04–064, HHSN271201300071C, RAG052527A) and from various national funding sources is gratefully acknowledged (see https://share-eric.eu/).

## Ethical approval

The SHARE study was approved by the Ethics Committee at the University of Mannheim (waves 1-4) and by the Ethics Council of the Max-Planck-Society (waves 5-8). Additionally, country-specific ethics committees or institutional review boards approved implementations of SHARE in the participating countries. All study participants provided informed consent.

## Availability of data and materials

Access to the SHARE data can be applied for with the SHARE Research Data Center (https://share-eric.eu/data). Code will be made available at https://github.com/kklot/SHARE/.

## Supplemental Information

### Survey questions related to the outcome

The following questions were used to collect the number of healthcare visits.

**Table S1:** Survey questions on number of healthcare visits.

| Wave | Question | Variable |
| --- | --- | --- |
| 1,2,4,5 | Now please think about the last 12 months. About how many times in total have you seen or talked to a medical doctor or qualified nurse about your health? Please exclude dentist visits and hospital stays, but include emergency room or outpatient clinic visits. | hc002_ |
| 6–8 | Now please think about the last 12 months. Since {month & year of last visit} about how many times in total have you seen or talked to a medical doctor or qualified/registered nurse about your health? Please exclude dentist visits and hospital stays, but include emergency room or outpatient clinic visits. | hc602_ |
| COVID II | Since last year/since last visit, did you go to a doctor's office or a medical facility other than a hospital? | caq120_ |

### Pre-pandemic trends in NHV

**Figure S1:**
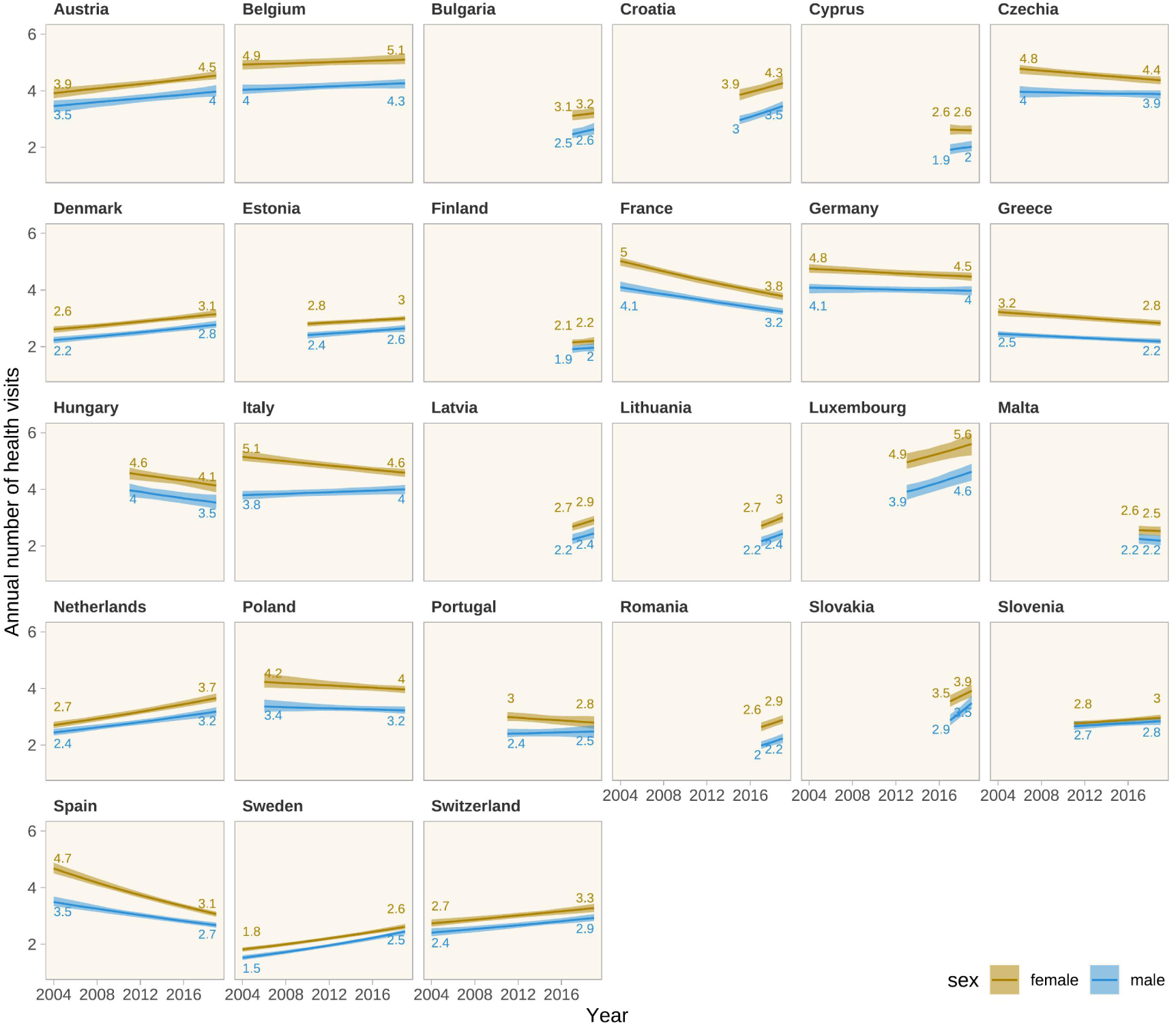
Posterior estimate of the annual rate of healthcare visits over time from 2004 to 2019. the line and its colour shade are the posterior median and 95% uncertainty interval. The estimates are conditioned on the reference group in the regression model: no comorbidities, an average age (approx. 75).

### Detailed estimates

**Table S2:** Estimate NHV by country and sex from 2004-2019.

| year | country | sex | 0.025quant | median | 0.975quant |
| --- | --- | --- | --- | --- | --- |
| 2004 | Austria | male | 3.244199 | 3.460106 | 3.662921 |
| 2005 | Austria | male | 3.284774 | 3.494889 | 3.680657 |
| 2006 | Austria | male | 3.325859 | 3.530927 | 3.704924 |
| 2007 | Austria | male | 3.379521 | 3.564859 | 3.721738 |
| 2008 | Austria | male | 3.425780 | 3.601352 | 3.740787 |
| 2009 | Austria | male | 3.464312 | 3.635793 | 3.764271 |
| 2010 | Austria | male | 3.503291 | 3.667981 | 3.795868 |
| 2011 | Austria | male | 3.551597 | 3.698551 | 3.832392 |
| 2012 | Austria | male | 3.594402 | 3.729973 | 3.870946 |
| 2013 | Austria | male | 3.633689 | 3.765922 | 3.905992 |
| 2014 | Austria | male | 3.665187 | 3.799657 | 3.947811 |
| 2015 | Austria | male | 3.694690 | 3.828295 | 3.994102 |
| 2016 | Austria | male | 3.730863 | 3.860150 | 4.041881 |
| 2017 | Austria | male | 3.762956 | 3.899215 | 4.082895 |
| 2018 | Austria | male | 3.782890 | 3.937422 | 4.138312 |
| 2019 | Austria | male | 3.800858 | 3.970442 | 4.201758 |
| 2020 | Austria | male | 3.823528 | 4.011040 | 4.259771 |
| 2021 | Austria | male | 3.847863 | 4.045841 | 4.321583 |
| 2022 | Austria | male | 3.867877 | 4.084259 | 4.384417 |
| 2017 | Bulgaria | male | 2.324126 | 2.464562 | 2.646342 |
| 2018 | Bulgaria | male | 2.404520 | 2.554225 | 2.722185 |
| 2019 | Bulgaria | male | 2.452132 | 2.643808 | 2.863810 |
| 2020 | Bulgaria | male | 2.481193 | 2.733460 | 3.012805 |
| 2021 | Bulgaria | male | 2.498934 | 2.817244 | 3.185286 |
| 2022 | Bulgaria | male | 2.516650 | 2.904888 | 3.395005 |
| 2004 | Belgium | male | 3.883873 | 4.034797 | 4.216610 |
| 2005 | Belgium | male | 3.909189 | 4.049865 | 4.215877 |
| 2006 | Belgium | male | 3.931470 | 4.060354 | 4.220124 |
| 2007 | Belgium | male | 3.952495 | 4.074560 | 4.233116 |
| 2008 | Belgium | male | 3.973634 | 4.088359 | 4.241866 |
| 2009 | Belgium | male | 3.995887 | 4.105081 | 4.249627 |
| 2010 | Belgium | male | 4.020443 | 4.123086 | 4.258893 |
| 2011 | Belgium | male | 4.046720 | 4.139696 | 4.272074 |
| 2012 | Belgium | male | 4.063808 | 4.154753 | 4.291558 |

Table S2: Estimate NHV by country and sex from 2004-2019 (*continued*)
| year | country | sex | 0.025quant | median | 0.975quant |
| --- | --- | --- | --- | --- | --- |
| 2013 | Belgium | male | 4.075209 | 4.171062 | 4.310870 |
| 2014 | Belgium | male | 4.082016 | 4.186349 | 4.325396 |
| 2015 | Belgium | male | 4.086260 | 4.202507 | 4.338838 |
| 2016 | Belgium | male | 4.085840 | 4.220226 | 4.354893 |
| 2017 | Belgium | male | 4.084775 | 4.236993 | 4.372816 |
| 2018 | Belgium | male | 4.083713 | 4.255554 | 4.393480 |
| 2019 | Belgium | male | 4.084788 | 4.263544 | 4.412957 |
| 2020 | Belgium | male | 4.085892 | 4.280969 | 4.436063 |
| 2021 | Belgium | male | 4.086998 | 4.297879 | 4.462347 |
| 2022 | Belgium | male | 4.088104 | 4.316731 | 4.490359 |
| 2017 | Cyprus | male | 1.753463 | 1.918135 | 2.113676 |
| 2018 | Cyprus | male | 1.824660 | 1.977191 | 2.169034 |
| 2019 | Cyprus | male | 1.865503 | 2.020889 | 2.230358 |
| 2020 | Cyprus | male | 1.854917 | 2.060596 | 2.317932 |
| 2021 | Cyprus | male | 1.844275 | 2.101663 | 2.476660 |
| 2022 | Cyprus | male | 1.816015 | 2.158657 | 2.627885 |
| 2006 | Czechia | male | 3.756379 | 3.965727 | 4.164976 |
| 2007 | Czechia | male | 3.770497 | 3.959211 | 4.134229 |
| 2008 | Czechia | male | 3.775132 | 3.951706 | 4.113152 |
| 2009 | Czechia | male | 3.781073 | 3.947550 | 4.093206 |
| 2010 | Czechia | male | 3.795763 | 3.935216 | 4.073388 |
| 2011 | Czechia | male | 3.802267 | 3.925542 | 4.058503 |
| 2012 | Czechia | male | 3.808273 | 3.916208 | 4.043924 |
| 2013 | Czechia | male | 3.807268 | 3.904517 | 4.032663 |
| 2014 | Czechia | male | 3.800561 | 3.897899 | 4.026677 |
| 2015 | Czechia | male | 3.796258 | 3.898441 | 4.021133 |
| 2016 | Czechia | male | 3.782812 | 3.895919 | 4.023218 |
| 2017 | Czechia | male | 3.763977 | 3.894924 | 4.021260 |
| 2018 | Czechia | male | 3.741245 | 3.892757 | 4.015659 |
| 2019 | Czechia | male | 3.717177 | 3.883397 | 4.015749 |
| 2020 | Czechia | male | 3.687792 | 3.878948 | 4.016611 |
| 2021 | Czechia | male | 3.661408 | 3.869957 | 4.017663 |
| 2022 | Czechia | male | 3.645380 | 3.862802 | 4.021453 |
| 2004 | Switzerland | male | 2.285286 | 2.411575 | 2.567788 |
| 2005 | Switzerland | male | 2.322030 | 2.443835 | 2.585608 |
| 2006 | Switzerland | male | 2.356833 | 2.475897 | 2.603658 |

*(Continued on Next Page...)*

Table S2: Estimate NHV by country and sex from 2004-2019 (*continued*)
| year | country | sex | 0.025quant | median | 0.975quant |
| --- | --- | --- | --- | --- | --- |
| 2007 | Switzerland | male | 2.392161 | 2.508115 | 2.622516 |
| 2008 | Switzerland | male | 2.428021 | 2.542401 | 2.649753 |
| 2009 | Switzerland | male | 2.464422 | 2.573775 | 2.679486 |
| 2010 | Switzerland | male | 2.502315 | 2.608691 | 2.709556 |
| 2011 | Switzerland | male | 2.542152 | 2.639242 | 2.740295 |
| 2012 | Switzerland | male | 2.585467 | 2.674719 | 2.771421 |
| 2013 | Switzerland | male | 2.628621 | 2.706673 | 2.807531 |
| 2014 | Switzerland | male | 2.667996 | 2.740811 | 2.848302 |
| 2015 | Switzerland | male | 2.702294 | 2.778900 | 2.886063 |
| 2016 | Switzerland | male | 2.723814 | 2.815149 | 2.924014 |
| 2017 | Switzerland | male | 2.752536 | 2.858882 | 2.971912 |
| 2018 | Switzerland | male | 2.788323 | 2.895651 | 3.020335 |
| 2019 | Switzerland | male | 2.823681 | 2.930536 | 3.069547 |
| 2020 | Switzerland | male | 2.859364 | 2.971176 | 3.119562 |
| 2021 | Switzerland | male | 2.895475 | 3.016268 | 3.170393 |
| 2022 | Switzerland | male | 2.925196 | 3.052734 | 3.222053 |
| 2004 | Germany | male | 3.880529 | 4.083841 | 4.217659 |
| 2005 | Germany | male | 3.891122 | 4.078301 | 4.200999 |
| 2006 | Germany | male | 3.899053 | 4.070844 | 4.186233 |
| 2007 | Germany | male | 3.900183 | 4.062363 | 4.172630 |
| 2008 | Germany | male | 3.901369 | 4.052025 | 4.160039 |
| 2009 | Germany | male | 3.912579 | 4.041371 | 4.147488 |
| 2010 | Germany | male | 3.917304 | 4.030796 | 4.136068 |
| 2011 | Germany | male | 3.919616 | 4.024849 | 4.128607 |
| 2012 | Germany | male | 3.911728 | 4.014345 | 4.125191 |
| 2013 | Germany | male | 3.905226 | 4.005981 | 4.115022 |
| 2014 | Germany | male | 3.892991 | 4.000654 | 4.108248 |
| 2015 | Germany | male | 3.879362 | 3.998325 | 4.106003 |
| 2016 | Germany | male | 3.863147 | 3.998550 | 4.107122 |
| 2017 | Germany | male | 3.846830 | 3.991262 | 4.113633 |
| 2018 | Germany | male | 3.827880 | 3.983048 | 4.125389 |
| 2019 | Germany | male | 3.810506 | 3.976494 | 4.134874 |
| 2020 | Germany | male | 3.794603 | 3.972071 | 4.146742 |
| 2021 | Germany | male | 3.772015 | 3.964498 | 4.158903 |
| 2022 | Germany | male | 3.748775 | 3.960792 | 4.171105 |
| 2004 | Denmark | male | 2.113838 | 2.229624 | 2.344852 |

*(Continued on Next Page...)*

Table S2: Estimate NHV by country and sex from 2004-2019 (*continued*)
| year | country | sex | 0.025quant | median | 0.975quant |
| --- | --- | --- | --- | --- | --- |
| 2005 | Denmark | male | 2.155025 | 2.261982 | 2.370705 |
| 2006 | Denmark | male | 2.196922 | 2.294652 | 2.400002 |
| 2007 | Denmark | male | 2.234987 | 2.328285 | 2.429858 |
| 2008 | Denmark | male | 2.273205 | 2.360738 | 2.457329 |
| 2009 | Denmark | male | 2.308850 | 2.396298 | 2.486311 |
| 2010 | Denmark | male | 2.339886 | 2.432487 | 2.517746 |
| 2011 | Denmark | male | 2.371347 | 2.468042 | 2.549649 |
| 2012 | Denmark | male | 2.408790 | 2.506387 | 2.582297 |
| 2013 | Denmark | male | 2.451103 | 2.543085 | 2.623545 |
| 2014 | Denmark | male | 2.492376 | 2.581734 | 2.664210 |
| 2015 | Denmark | male | 2.526747 | 2.619157 | 2.709997 |
| 2016 | Denmark | male | 2.559962 | 2.656521 | 2.759732 |
| 2017 | Denmark | male | 2.596059 | 2.696212 | 2.810565 |
| 2018 | Denmark | male | 2.632531 | 2.737158 | 2.861284 |
| 2019 | Denmark | male | 2.669349 | 2.774810 | 2.912520 |
| 2020 | Denmark | male | 2.703145 | 2.815101 | 2.964678 |
| 2021 | Denmark | male | 2.735676 | 2.857313 | 3.017773 |
| 2022 | Denmark | male | 2.768526 | 2.898941 | 3.071823 |
| 2010 | Estonia | male | 2.285376 | 2.407619 | 2.515785 |
| 2011 | Estonia | male | 2.321620 | 2.435062 | 2.531984 |
| 2012 | Estonia | male | 2.354188 | 2.461547 | 2.554775 |
| 2013 | Estonia | male | 2.387216 | 2.485227 | 2.576618 |
| 2014 | Estonia | male | 2.420712 | 2.508629 | 2.607167 |
| 2015 | Estonia | male | 2.451066 | 2.532520 | 2.639991 |
| 2016 | Estonia | male | 2.482253 | 2.560670 | 2.662277 |
| 2017 | Estonia | male | 2.500171 | 2.588182 | 2.691715 |
| 2018 | Estonia | male | 2.512945 | 2.617084 | 2.728164 |
| 2019 | Estonia | male | 2.521022 | 2.649383 | 2.765736 |
| 2020 | Estonia | male | 2.533602 | 2.680576 | 2.809078 |
| 2021 | Estonia | male | 2.546200 | 2.709136 | 2.856455 |
| 2022 | Estonia | male | 2.558527 | 2.742129 | 2.904632 |
| 2004 | Spain | male | 3.347348 | 3.498001 | 3.688898 |
| 2005 | Spain | male | 3.297239 | 3.432552 | 3.613407 |
| 2006 | Spain | male | 3.246230 | 3.369188 | 3.537009 |
| 2007 | Spain | male | 3.194815 | 3.311524 | 3.459134 |
| 2008 | Spain | male | 3.142192 | 3.259413 | 3.377703 |

*(Continued on Next Page...)*

Table S2: Estimate NHV by country and sex from 2004-2019 (*continued*)
| year | country | sex | 0.025quant | median | 0.975quant |
| --- | --- | --- | --- | --- | --- |
| 2009 | Spain | male | 3.090435 | 3.200742 | 3.300218 |
| 2010 | Spain | male | 3.039625 | 3.144317 | 3.231704 |
| 2011 | Spain | male | 2.989466 | 3.088112 | 3.171840 |
| 2012 | Spain | male | 2.939894 | 3.033418 | 3.117152 |
| 2013 | Spain | male | 2.889127 | 2.982913 | 3.061398 |
| 2014 | Spain | male | 2.839237 | 2.925885 | 3.008013 |
| 2015 | Spain | male | 2.790210 | 2.870999 | 2.951013 |
| 2016 | Spain | male | 2.740510 | 2.822379 | 2.903303 |
| 2017 | Spain | male | 2.690462 | 2.771261 | 2.858577 |
| 2018 | Spain | male | 2.641328 | 2.722688 | 2.813158 |
| 2019 | Spain | male | 2.586018 | 2.674989 | 2.769134 |
| 2020 | Spain | male | 2.528921 | 2.628114 | 2.727162 |
| 2021 | Spain | male | 2.475114 | 2.581005 | 2.686196 |
| 2022 | Spain | male | 2.422000 | 2.534916 | 2.645852 |
| 2004 | France | male | 3.954110 | 4.101470 | 4.299594 |
| 2005 | France | male | 3.902900 | 4.035913 | 4.220847 |
| 2006 | France | male | 3.852355 | 3.969498 | 4.143909 |
| 2007 | France | male | 3.799137 | 3.908429 | 4.070009 |
| 2008 | France | male | 3.750196 | 3.854564 | 4.000267 |
| 2009 | France | male | 3.697364 | 3.798958 | 3.929466 |
| 2010 | France | male | 3.645561 | 3.741282 | 3.860323 |
| 2011 | France | male | 3.589038 | 3.685064 | 3.792560 |
| 2012 | France | male | 3.529655 | 3.625250 | 3.725994 |
| 2013 | France | male | 3.469672 | 3.564048 | 3.667587 |
| 2014 | France | male | 3.410708 | 3.510265 | 3.618347 |
| 2015 | France | male | 3.352746 | 3.452330 | 3.568869 |
| 2016 | France | male | 3.295770 | 3.393085 | 3.518156 |
| 2017 | France | male | 3.239762 | 3.342080 | 3.460407 |
| 2018 | France | male | 3.184701 | 3.286363 | 3.407942 |
| 2019 | France | male | 3.130243 | 3.235844 | 3.357987 |
| 2020 | France | male | 3.075435 | 3.182166 | 3.309031 |
| 2021 | France | male | 3.022239 | 3.133501 | 3.263499 |
| 2022 | France | male | 2.968287 | 3.086178 | 3.221732 |
| 2017 | Finland | male | 1.776112 | 1.912624 | 2.029705 |
| 2018 | Finland | male | 1.828131 | 1.937079 | 2.073979 |
| 2019 | Finland | male | 1.838486 | 1.963458 | 2.120321 |

*(Continued on Next Page...)*

Table S2: Estimate NHV by country and sex from 2004-2019 (*continued*)
| year | country | sex | 0.025quant | median | 0.975quant |
| --- | --- | --- | --- | --- | --- |
| 2020 | Finland | male | 1.821745 | 1.989490 | 2.217819 |
| 2021 | Finland | male | 1.797792 | 2.022566 | 2.329718 |
| 2022 | Finland | male | 1.762212 | 2.061379 | 2.437676 |
| 2004 | Greece | male | 2.336756 | 2.455857 | 2.546893 |
| 2005 | Greece | male | 2.327260 | 2.437612 | 2.520586 |
| 2006 | Greece | male | 2.320755 | 2.420404 | 2.496679 |
| 2007 | Greece | male | 2.310035 | 2.401591 | 2.473771 |
| 2008 | Greece | male | 2.295108 | 2.383327 | 2.451333 |
| 2009 | Greece | male | 2.282930 | 2.364572 | 2.431897 |
| 2010 | Greece | male | 2.270677 | 2.345659 | 2.414553 |
| 2011 | Greece | male | 2.255879 | 2.325607 | 2.397333 |
| 2012 | Greece | male | 2.239666 | 2.306747 | 2.380236 |
| 2013 | Greece | male | 2.222699 | 2.288724 | 2.366024 |
| 2014 | Greece | male | 2.203847 | 2.269812 | 2.351651 |
| 2015 | Greece | male | 2.184191 | 2.255534 | 2.330872 |
| 2016 | Greece | male | 2.163298 | 2.237645 | 2.318205 |
| 2017 | Greece | male | 2.142575 | 2.221655 | 2.305785 |
| 2018 | Greece | male | 2.121995 | 2.202893 | 2.293656 |
| 2019 | Greece | male | 2.101430 | 2.183801 | 2.281607 |
| 2020 | Greece | male | 2.080759 | 2.166986 | 2.271962 |
| 2021 | Greece | male | 2.060125 | 2.148391 | 2.260097 |
| 2022 | Greece | male | 2.039696 | 2.129329 | 2.249710 |
| 2015 | Croatia | male | 2.818899 | 2.963210 | 3.125338 |
| 2016 | Croatia | male | 2.931890 | 3.079602 | 3.211470 |
| 2017 | Croatia | male | 3.061638 | 3.202042 | 3.325886 |
| 2018 | Croatia | male | 3.162173 | 3.327052 | 3.474513 |
| 2019 | Croatia | male | 3.266322 | 3.459496 | 3.634036 |
| 2020 | Croatia | male | 3.348277 | 3.588136 | 3.809929 |
| 2021 | Croatia | male | 3.420714 | 3.723385 | 3.987053 |
| 2022 | Croatia | male | 3.504930 | 3.868867 | 4.182924 |
| 2011 | Hungary | male | 3.737053 | 3.961451 | 4.198961 |
| 2012 | Hungary | male | 3.694354 | 3.912721 | 4.111955 |
| 2013 | Hungary | male | 3.655128 | 3.845653 | 4.035131 |
| 2014 | Hungary | male | 3.603356 | 3.786226 | 3.994130 |
| 2015 | Hungary | male | 3.547816 | 3.738637 | 3.937473 |
| 2016 | Hungary | male | 3.493081 | 3.678122 | 3.891035 |

*(Continued on Next Page...)*

Table S2: Estimate NHV by country and sex from 2004-2019 (*continued*)
| year | country | sex | 0.025quant | median | 0.975quant |
| --- | --- | --- | --- | --- | --- |
| 2017 | Hungary | male | 3.433541 | 3.628272 | 3.849194 |
| 2018 | Hungary | male | 3.377104 | 3.581787 | 3.819606 |
| 2019 | Hungary | male | 3.310496 | 3.527093 | 3.811497 |
| 2020 | Hungary | male | 3.239111 | 3.476036 | 3.801276 |
| 2021 | Hungary | male | 3.159718 | 3.423566 | 3.781077 |
| 2022 | Hungary | male | 3.082273 | 3.373404 | 3.761957 |
| 2004 | Italy | male | 3.638012 | 3.792002 | 3.940272 |
| 2005 | Italy | male | 3.655998 | 3.804094 | 3.945304 |
| 2006 | Italy | male | 3.677391 | 3.817871 | 3.950347 |
| 2007 | Italy | male | 3.701692 | 3.826553 | 3.955567 |
| 2008 | Italy | male | 3.721287 | 3.836572 | 3.966581 |
| 2009 | Italy | male | 3.742348 | 3.855299 | 3.981572 |
| 2010 | Italy | male | 3.755036 | 3.870574 | 3.993130 |
| 2011 | Italy | male | 3.762534 | 3.881574 | 4.003240 |
| 2012 | Italy | male | 3.774827 | 3.893342 | 4.013815 |
| 2013 | Italy | male | 3.785281 | 3.909547 | 4.033153 |
| 2014 | Italy | male | 3.793788 | 3.923897 | 4.053603 |
| 2015 | Italy | male | 3.802878 | 3.939104 | 4.068472 |
| 2016 | Italy | male | 3.814293 | 3.951878 | 4.083120 |
| 2017 | Italy | male | 3.819651 | 3.966996 | 4.105077 |
| 2018 | Italy | male | 3.820580 | 3.983181 | 4.129362 |
| 2019 | Italy | male | 3.821325 | 3.998240 | 4.153798 |
| 2020 | Italy | male | 3.822070 | 4.016486 | 4.178386 |
| 2021 | Italy | male | 3.822815 | 4.029089 | 4.203538 |
| 2022 | Italy | male | 3.823559 | 4.043919 | 4.228974 |
| 2017 | Lithuania | male | 1.966791 | 2.161937 | 2.320051 |
| 2018 | Lithuania | male | 2.119839 | 2.294761 | 2.443565 |
| 2019 | Lithuania | male | 2.257654 | 2.431049 | 2.598693 |
| 2020 | Lithuania | male | 2.367347 | 2.583298 | 2.808093 |
| 2021 | Lithuania | male | 2.480978 | 2.745333 | 3.056385 |
| 2022 | Lithuania | male | 2.573293 | 2.914560 | 3.359269 |
| 2013 | Luxembourg | male | 3.629288 | 3.914485 | 4.153103 |
| 2014 | Luxembourg | male | 3.750787 | 4.008530 | 4.237594 |
| 2015 | Luxembourg | male | 3.863161 | 4.127877 | 4.337275 |
| 2016 | Luxembourg | male | 3.978641 | 4.238819 | 4.469403 |
| 2017 | Luxembourg | male | 4.095715 | 4.367421 | 4.622146 |

*(Continued on Next Page...)*

Table S2: Estimate NHV by country and sex from 2004-2019 (*continued*)
| year | country | sex | 0.025quant | median | 0.975quant |
| --- | --- | --- | --- | --- | --- |
| 2018 | Luxembourg | male | 4.199950 | 4.488704 | 4.750377 |
| 2019 | Luxembourg | male | 4.303494 | 4.618392 | 4.900402 |
| 2020 | Luxembourg | male | 4.399428 | 4.755745 | 5.124929 |
| 2021 | Luxembourg | male | 4.480262 | 4.896041 | 5.323812 |
| 2022 | Luxembourg | male | 4.571260 | 5.014299 | 5.529228 |
| 2017 | Latvia | male | 2.057815 | 2.223410 | 2.417324 |
| 2018 | Latvia | male | 2.183627 | 2.330396 | 2.513813 |
| 2019 | Latvia | male | 2.256063 | 2.440169 | 2.665699 |
| 2020 | Latvia | male | 2.291690 | 2.559015 | 2.888447 |
| 2021 | Latvia | male | 2.315060 | 2.680334 | 3.102717 |
| 2022 | Latvia | male | 2.339417 | 2.810347 | 3.348298 |
| 2017 | Malta | male | 2.069205 | 2.245738 | 2.423255 |
| 2018 | Malta | male | 2.053578 | 2.213989 | 2.392156 |
| 2019 | Malta | male | 2.010926 | 2.183875 | 2.366208 |
| 2020 | Malta | male | 1.932824 | 2.161301 | 2.377905 |
| 2021 | Malta | male | 1.869101 | 2.129763 | 2.428568 |
| 2022 | Malta | male | 1.795742 | 2.104780 | 2.473381 |
| 2004 | Netherlands | male | 2.341247 | 2.445362 | 2.574740 |
| 2005 | Netherlands | male | 2.385838 | 2.488632 | 2.612018 |
| 2006 | Netherlands | male | 2.430836 | 2.532519 | 2.655612 |
| 2007 | Netherlands | male | 2.485407 | 2.576640 | 2.694141 |
| 2008 | Netherlands | male | 2.539309 | 2.624417 | 2.731684 |
| 2009 | Netherlands | male | 2.589852 | 2.670062 | 2.771262 |
| 2010 | Netherlands | male | 2.636201 | 2.715317 | 2.813063 |
| 2011 | Netherlands | male | 2.679952 | 2.761771 | 2.856186 |
| 2012 | Netherlands | male | 2.720350 | 2.812735 | 2.902204 |
| 2013 | Netherlands | male | 2.763426 | 2.860812 | 2.950651 |
| 2014 | Netherlands | male | 2.807184 | 2.913119 | 3.008147 |
| 2015 | Netherlands | male | 2.851116 | 2.969082 | 3.067172 |
| 2016 | Netherlands | male | 2.891873 | 3.022309 | 3.129340 |
| 2017 | Netherlands | male | 2.931165 | 3.073998 | 3.194306 |
| 2018 | Netherlands | male | 2.971659 | 3.125741 | 3.263563 |
| 2019 | Netherlands | male | 3.018083 | 3.183243 | 3.335922 |
| 2020 | Netherlands | male | 3.065121 | 3.238432 | 3.415960 |
| 2021 | Netherlands | male | 3.112610 | 3.293271 | 3.497872 |
| 2022 | Netherlands | male | 3.159560 | 3.348987 | 3.570559 |

*(Continued on Next Page...)*

Table S2: Estimate NHV by country and sex from 2004-2019 (*continued*)
| year | country | sex | 0.025quant | median | 0.975quant |
| --- | --- | --- | --- | --- | --- |
| 2006 | Poland | male | 3.177556 | 3.363993 | 3.611440 |
| 2007 | Poland | male | 3.185707 | 3.351467 | 3.578940 |
| 2008 | Poland | male | 3.189846 | 3.337737 | 3.546882 |
| 2009 | Poland | male | 3.193583 | 3.327522 | 3.517528 |
| 2010 | Poland | male | 3.200949 | 3.314843 | 3.484600 |
| 2011 | Poland | male | 3.207288 | 3.303818 | 3.455098 |
| 2012 | Poland | male | 3.207839 | 3.300330 | 3.431305 |
| 2013 | Poland | male | 3.203941 | 3.288313 | 3.408860 |
| 2014 | Poland | male | 3.199674 | 3.279612 | 3.395817 |
| 2015 | Poland | male | 3.189393 | 3.267826 | 3.385257 |
| 2016 | Poland | male | 3.172290 | 3.263330 | 3.375812 |
| 2017 | Poland | male | 3.153659 | 3.248354 | 3.375024 |
| 2018 | Poland | male | 3.131477 | 3.234993 | 3.368743 |
| 2019 | Poland | male | 3.105822 | 3.225807 | 3.366856 |
| 2020 | Poland | male | 3.080379 | 3.216849 | 3.369025 |
| 2021 | Poland | male | 3.054464 | 3.208562 | 3.374845 |
| 2022 | Poland | male | 3.025946 | 3.198320 | 3.382920 |
| 2011 | Portugal | male | 2.272455 | 2.401393 | 2.579618 |
| 2012 | Portugal | male | 2.302620 | 2.407835 | 2.569256 |
| 2013 | Portugal | male | 2.307798 | 2.412541 | 2.566413 |
| 2014 | Portugal | male | 2.308548 | 2.429650 | 2.564225 |
| 2015 | Portugal | male | 2.307666 | 2.439369 | 2.576635 |
| 2016 | Portugal | male | 2.302540 | 2.450754 | 2.603310 |
| 2017 | Portugal | male | 2.286131 | 2.460403 | 2.633796 |
| 2018 | Portugal | male | 2.267427 | 2.469431 | 2.664843 |
| 2019 | Portugal | male | 2.245716 | 2.478152 | 2.698444 |
| 2020 | Portugal | male | 2.216033 | 2.490652 | 2.747678 |
| 2021 | Portugal | male | 2.182451 | 2.508151 | 2.805644 |
| 2022 | Portugal | male | 2.156685 | 2.522416 | 2.854331 |
| 2017 | Romania | male | 1.865088 | 1.984371 | 2.130991 |
| 2018 | Romania | male | 1.989411 | 2.103079 | 2.256252 |
| 2019 | Romania | male | 2.068890 | 2.233734 | 2.402204 |
| 2020 | Romania | male | 2.141704 | 2.364406 | 2.579603 |
| 2021 | Romania | male | 2.213788 | 2.505229 | 2.785167 |
| 2022 | Romania | male | 2.303017 | 2.659305 | 3.013512 |
| 2004 | Sweden | male | 1.448451 | 1.523429 | 1.611074 |

*(Continued on Next Page...)*

Table S2: Estimate NHV by country and sex from 2004-2019 (*continued*)
| year | country | sex | 0.025quant | median | 0.975quant |
| --- | --- | --- | --- | --- | --- |
| 2005 | Sweden | male | 1.498871 | 1.572926 | 1.656019 |
| 2006 | Sweden | male | 1.551047 | 1.623066 | 1.702220 |
| 2007 | Sweden | male | 1.604913 | 1.674941 | 1.748121 |
| 2008 | Sweden | male | 1.660805 | 1.728399 | 1.797049 |
| 2009 | Sweden | male | 1.720712 | 1.783870 | 1.847507 |
| 2010 | Sweden | male | 1.780412 | 1.841917 | 1.899388 |
| 2011 | Sweden | male | 1.840181 | 1.901901 | 1.953293 |
| 2012 | Sweden | male | 1.897795 | 1.962628 | 2.012525 |
| 2013 | Sweden | male | 1.958306 | 2.025201 | 2.078350 |
| 2014 | Sweden | male | 2.016908 | 2.091179 | 2.147002 |
| 2015 | Sweden | male | 2.081239 | 2.158060 | 2.211285 |
| 2016 | Sweden | male | 2.146044 | 2.227174 | 2.280271 |
| 2017 | Sweden | male | 2.212870 | 2.301468 | 2.355540 |
| 2018 | Sweden | male | 2.280838 | 2.375196 | 2.436340 |
| 2019 | Sweden | male | 2.346240 | 2.451948 | 2.518284 |
| 2020 | Sweden | male | 2.412804 | 2.531435 | 2.607695 |
| 2021 | Sweden | male | 2.481146 | 2.615391 | 2.698335 |
| 2022 | Sweden | male | 2.552238 | 2.700723 | 2.793456 |
| 2011 | Slovenia | male | 2.539055 | 2.662643 | 2.835017 |
| 2012 | Slovenia | male | 2.574255 | 2.688518 | 2.837687 |
| 2013 | Slovenia | male | 2.609459 | 2.707287 | 2.847667 |
| 2014 | Slovenia | male | 2.642271 | 2.735856 | 2.848931 |
| 2015 | Slovenia | male | 2.672911 | 2.754136 | 2.855295 |
| 2016 | Slovenia | male | 2.688128 | 2.775179 | 2.879561 |
| 2017 | Slovenia | male | 2.700513 | 2.790653 | 2.903197 |
| 2018 | Slovenia | male | 2.709785 | 2.821380 | 2.934738 |
| 2019 | Slovenia | male | 2.717089 | 2.842528 | 2.981454 |
| 2020 | Slovenia | male | 2.724286 | 2.866570 | 3.027800 |
| 2021 | Slovenia | male | 2.727186 | 2.881719 | 3.075243 |
| 2022 | Slovenia | male | 2.722732 | 2.899404 | 3.123445 |
| 2017 | Slovakia | male | 2.691845 | 2.876603 | 3.078890 |
| 2018 | Slovakia | male | 2.995293 | 3.166089 | 3.366906 |
| 2019 | Slovakia | male | 3.271768 | 3.484018 | 3.741000 |
| 2020 | Slovakia | male | 3.551516 | 3.842132 | 4.172829 |
| 2021 | Slovakia | male | 3.829498 | 4.235402 | 4.704391 |
| 2022 | Slovakia | male | 4.128872 | 4.680766 | 5.314641 |

*(Continued on Next Page...)*

Table S2: Estimate NHV by country and sex from 2004-2019 (*continued*)
| year | country | sex | 0.025quant | median | 0.975quant |
| --- | --- | --- | --- | --- | --- |
| 2004 | Austria | female | 3.735494 | 3.915671 | 4.110365 |
| 2005 | Austria | female | 3.784077 | 3.957955 | 4.132839 |
| 2006 | Austria | female | 3.833354 | 4.001247 | 4.156065 |
| 2007 | Austria | female | 3.883275 | 4.045384 | 4.186932 |
| 2008 | Austria | female | 3.933680 | 4.082222 | 4.220465 |
| 2009 | Austria | female | 3.988576 | 4.121603 | 4.254267 |
| 2010 | Austria | female | 4.040944 | 4.161906 | 4.288339 |
| 2011 | Austria | female | 4.087868 | 4.199689 | 4.322685 |
| 2012 | Austria | female | 4.132165 | 4.237815 | 4.357333 |
| 2013 | Austria | female | 4.184429 | 4.276999 | 4.389360 |
| 2014 | Austria | female | 4.234605 | 4.316763 | 4.429903 |
| 2015 | Austria | female | 4.275584 | 4.358605 | 4.480788 |
| 2016 | Austria | female | 4.310204 | 4.403287 | 4.532076 |
| 2017 | Austria | female | 4.345091 | 4.449014 | 4.585876 |
| 2018 | Austria | female | 4.376074 | 4.496176 | 4.640846 |
| 2019 | Austria | female | 4.407278 | 4.538810 | 4.696478 |
| 2020 | Austria | female | 4.435362 | 4.583822 | 4.752780 |
| 2021 | Austria | female | 4.463632 | 4.634616 | 4.809759 |
| 2022 | Austria | female | 4.499031 | 4.684298 | 4.867805 |
| 2017 | Bulgaria | female | 2.955254 | 3.118614 | 3.321842 |
| 2018 | Bulgaria | female | 2.994959 | 3.162940 | 3.339415 |
| 2019 | Bulgaria | female | 3.033511 | 3.212301 | 3.395708 |
| 2020 | Bulgaria | female | 3.045902 | 3.247493 | 3.457382 |
| 2021 | Bulgaria | female | 3.071607 | 3.303859 | 3.534979 |
| 2022 | Bulgaria | female | 3.064799 | 3.345219 | 3.629517 |
| 2004 | Belgium | female | 4.753337 | 4.928705 | 5.084123 |
| 2005 | Belgium | female | 4.768259 | 4.939547 | 5.088362 |
| 2006 | Belgium | female | 4.784249 | 4.949718 | 5.087760 |
| 2007 | Belgium | female | 4.799884 | 4.960388 | 5.090092 |
| 2008 | Belgium | female | 4.822057 | 4.968367 | 5.097161 |
| 2009 | Belgium | female | 4.844422 | 4.979019 | 5.112787 |
| 2010 | Belgium | female | 4.864963 | 4.989918 | 5.127725 |
| 2011 | Belgium | female | 4.885593 | 5.000024 | 5.138914 |
| 2012 | Belgium | female | 4.905554 | 5.011070 | 5.153849 |
| 2013 | Belgium | female | 4.910867 | 5.024671 | 5.169136 |
| 2014 | Belgium | female | 4.915651 | 5.041120 | 5.186393 |

*(Continued on Next Page...)*

Table S2: Estimate NHV by country and sex from 2004-2019 (*continued*)
| year | country | sex | 0.025quant | median | 0.975quant |
| --- | --- | --- | --- | --- | --- |
| 2015 | Belgium | female | 4.925226 | 5.050290 | 5.206600 |
| 2016 | Belgium | female | 4.931914 | 5.063828 | 5.226887 |
| 2017 | Belgium | female | 4.935126 | 5.073770 | 5.247254 |
| 2018 | Belgium | female | 4.938343 | 5.084815 | 5.267701 |
| 2019 | Belgium | female | 4.941566 | 5.101371 | 5.288228 |
| 2020 | Belgium | female | 4.944794 | 5.115536 | 5.308836 |
| 2021 | Belgium | female | 4.949297 | 5.128701 | 5.329524 |
| 2022 | Belgium | female | 4.954104 | 5.137231 | 5.349836 |
| 2017 | Cyprus | female | 2.446475 | 2.630950 | 2.813923 |
| 2018 | Cyprus | female | 2.470842 | 2.615918 | 2.770624 |
| 2019 | Cyprus | female | 2.459626 | 2.605609 | 2.777563 |
| 2020 | Cyprus | female | 2.399854 | 2.600710 | 2.790719 |
| 2021 | Cyprus | female | 2.336283 | 2.591249 | 2.799189 |
| 2022 | Cyprus | female | 2.274287 | 2.598293 | 2.863892 |
| 2006 | Czechia | female | 4.599881 | 4.777091 | 4.913460 |
| 2007 | Czechia | female | 4.574558 | 4.745225 | 4.870537 |
| 2008 | Czechia | female | 4.549155 | 4.712213 | 4.827989 |
| 2009 | Czechia | female | 4.523896 | 4.684113 | 4.787394 |
| 2010 | Czechia | female | 4.495609 | 4.653271 | 4.750305 |
| 2011 | Czechia | female | 4.471246 | 4.619495 | 4.718923 |
| 2012 | Czechia | female | 4.446398 | 4.588185 | 4.683594 |
| 2013 | Czechia | female | 4.421278 | 4.555365 | 4.651769 |
| 2014 | Czechia | female | 4.389729 | 4.524141 | 4.625032 |
| 2015 | Czechia | female | 4.359839 | 4.497901 | 4.597731 |
| 2016 | Czechia | female | 4.323843 | 4.467589 | 4.573183 |
| 2017 | Czechia | female | 4.286892 | 4.436518 | 4.549060 |
| 2018 | Czechia | female | 4.260820 | 4.404281 | 4.527169 |
| 2019 | Czechia | female | 4.234907 | 4.372992 | 4.509752 |
| 2020 | Czechia | female | 4.208398 | 4.342969 | 4.492409 |
| 2021 | Czechia | female | 4.170297 | 4.310345 | 4.475433 |
| 2022 | Czechia | female | 4.130555 | 4.280354 | 4.463742 |
| 2004 | Switzerland | female | 2.626681 | 2.739179 | 2.879000 |
| 2005 | Switzerland | female | 2.667778 | 2.771767 | 2.905889 |
| 2006 | Switzerland | female | 2.708207 | 2.805741 | 2.933115 |
| 2007 | Switzerland | female | 2.747922 | 2.841061 | 2.960691 |
| 2008 | Switzerland | female | 2.786662 | 2.873517 | 2.988528 |

*(Continued on Next Page...)*

Table S2: Estimate NHV by country and sex from 2004-2019 (*continued*)
| year | country | sex | 0.025quant | median | 0.975quant |
| --- | --- | --- | --- | --- | --- |
| 2009 | Switzerland | female | 2.827438 | 2.909365 | 3.016853 |
| 2010 | Switzerland | female | 2.860361 | 2.948323 | 3.048654 |
| 2011 | Switzerland | female | 2.895703 | 2.986844 | 3.080790 |
| 2012 | Switzerland | female | 2.933433 | 3.022768 | 3.114332 |
| 2013 | Switzerland | female | 2.965850 | 3.056185 | 3.147747 |
| 2014 | Switzerland | female | 2.997005 | 3.094638 | 3.180756 |
| 2015 | Switzerland | female | 3.026416 | 3.132675 | 3.226170 |
| 2016 | Switzerland | female | 3.053507 | 3.169798 | 3.278668 |
| 2017 | Switzerland | female | 3.081347 | 3.204209 | 3.321897 |
| 2018 | Switzerland | female | 3.110112 | 3.244129 | 3.375464 |
| 2019 | Switzerland | female | 3.139148 | 3.279264 | 3.431675 |
| 2020 | Switzerland | female | 3.168340 | 3.317385 | 3.491507 |
| 2021 | Switzerland | female | 3.195890 | 3.360054 | 3.552381 |
| 2022 | Switzerland | female | 3.224190 | 3.400590 | 3.614317 |
| 2004 | Germany | female | 4.608425 | 4.754839 | 4.916966 |
| 2005 | Germany | female | 4.599849 | 4.735299 | 4.886126 |
| 2006 | Germany | female | 4.586269 | 4.717227 | 4.857341 |
| 2007 | Germany | female | 4.572083 | 4.697857 | 4.833489 |
| 2008 | Germany | female | 4.557211 | 4.680821 | 4.809757 |
| 2009 | Germany | female | 4.545744 | 4.658635 | 4.783771 |
| 2010 | Germany | female | 4.530738 | 4.634310 | 4.758261 |
| 2011 | Germany | female | 4.513707 | 4.613086 | 4.734183 |
| 2012 | Germany | female | 4.490801 | 4.592418 | 4.712471 |
| 2013 | Germany | female | 4.467426 | 4.576897 | 4.693168 |
| 2014 | Germany | female | 4.444380 | 4.558130 | 4.676685 |
| 2015 | Germany | female | 4.422837 | 4.541738 | 4.668512 |
| 2016 | Germany | female | 4.404281 | 4.523478 | 4.654200 |
| 2017 | Germany | female | 4.382764 | 4.508930 | 4.638314 |
| 2018 | Germany | female | 4.358576 | 4.492731 | 4.629526 |
| 2019 | Germany | female | 4.327364 | 4.474504 | 4.616664 |
| 2020 | Germany | female | 4.295916 | 4.455082 | 4.603838 |
| 2021 | Germany | female | 4.264699 | 4.435746 | 4.591048 |
| 2022 | Germany | female | 4.237906 | 4.418943 | 4.578294 |
| 2004 | Denmark | female | 2.488565 | 2.605938 | 2.717544 |
| 2005 | Denmark | female | 2.526139 | 2.640767 | 2.740164 |
| 2006 | Denmark | female | 2.564281 | 2.671154 | 2.762975 |

*(Continued on Next Page...)*

Table S2: Estimate NHV by country and sex from 2004-2019 (*continued*)
| year | country | sex | 0.025quant | median | 0.975quant |
| --- | --- | --- | --- | --- | --- |
| 2007 | Denmark | female | 2.602998 | 2.704470 | 2.788135 |
| 2008 | Denmark | female | 2.642300 | 2.738037 | 2.818199 |
| 2009 | Denmark | female | 2.680643 | 2.774369 | 2.851143 |
| 2010 | Denmark | female | 2.720850 | 2.809468 | 2.882828 |
| 2011 | Denmark | female | 2.759447 | 2.845588 | 2.925323 |
| 2012 | Denmark | female | 2.801874 | 2.883952 | 2.965726 |
| 2013 | Denmark | female | 2.843085 | 2.918636 | 3.006261 |
| 2014 | Denmark | female | 2.881737 | 2.958353 | 3.044103 |
| 2015 | Denmark | female | 2.913538 | 2.991723 | 3.088167 |
| 2016 | Denmark | female | 2.945451 | 3.032236 | 3.134852 |
| 2017 | Denmark | female | 2.976872 | 3.072948 | 3.185511 |
| 2018 | Denmark | female | 3.009854 | 3.114324 | 3.237803 |
| 2019 | Denmark | female | 3.039793 | 3.149028 | 3.288694 |
| 2020 | Denmark | female | 3.064266 | 3.191208 | 3.337181 |
| 2021 | Denmark | female | 3.091046 | 3.231469 | 3.388680 |
| 2022 | Denmark | female | 3.121496 | 3.271005 | 3.441476 |
| 2010 | Estonia | female | 2.712585 | 2.807739 | 2.885302 |
| 2011 | Estonia | female | 2.740018 | 2.828649 | 2.903997 |
| 2012 | Estonia | female | 2.764124 | 2.849791 | 2.924152 |
| 2013 | Estonia | female | 2.783987 | 2.870728 | 2.943813 |
| 2014 | Estonia | female | 2.803679 | 2.893038 | 2.960213 |
| 2015 | Estonia | female | 2.829431 | 2.911736 | 2.977041 |
| 2016 | Estonia | female | 2.855434 | 2.934417 | 3.003339 |
| 2017 | Estonia | female | 2.872642 | 2.955193 | 3.026714 |
| 2018 | Estonia | female | 2.889775 | 2.977300 | 3.054309 |
| 2019 | Estonia | female | 2.911482 | 2.999718 | 3.082475 |
| 2020 | Estonia | female | 2.928030 | 3.022951 | 3.110941 |
| 2021 | Estonia | female | 2.941342 | 3.045296 | 3.139833 |
| 2022 | Estonia | female | 2.956208 | 3.069997 | 3.168887 |
| 2004 | Spain | female | 4.504645 | 4.679303 | 4.882818 |
| 2005 | Spain | female | 4.398208 | 4.548768 | 4.739438 |
| 2006 | Spain | female | 4.289840 | 4.423814 | 4.600748 |
| 2007 | Spain | female | 4.177624 | 4.299755 | 4.466166 |
| 2008 | Spain | female | 4.066658 | 4.180841 | 4.335046 |
| 2009 | Spain | female | 3.965286 | 4.062709 | 4.206594 |
| 2010 | Spain | female | 3.858072 | 3.952412 | 4.078485 |

*(Continued on Next Page...)*

Table S2: Estimate NHV by country and sex from 2004-2019 (*continued*)
| year | country | sex | 0.025quant | median | 0.975quant |
| --- | --- | --- | --- | --- | --- |
| 2011 | Spain | female | 3.748886 | 3.842315 | 3.958075 |
| 2012 | Spain | female | 3.642618 | 3.737187 | 3.843626 |
| 2013 | Spain | female | 3.545995 | 3.636059 | 3.730048 |
| 2014 | Spain | female | 3.441441 | 3.536817 | 3.621179 |
| 2015 | Spain | female | 3.344977 | 3.440848 | 3.516975 |
| 2016 | Spain | female | 3.251491 | 3.346616 | 3.420922 |
| 2017 | Spain | female | 3.156576 | 3.252368 | 3.328890 |
| 2018 | Spain | female | 3.063828 | 3.159841 | 3.241222 |
| 2019 | Spain | female | 2.975365 | 3.069551 | 3.157538 |
| 2020 | Spain | female | 2.889302 | 2.985033 | 3.076954 |
| 2021 | Spain | female | 2.805730 | 2.903955 | 2.998559 |
| 2022 | Spain | female | 2.723722 | 2.823784 | 2.922291 |
| 2004 | France | female | 4.860738 | 5.026449 | 5.154656 |
| 2005 | France | female | 4.780526 | 4.935088 | 5.053465 |
| 2006 | France | female | 4.703313 | 4.843147 | 4.954260 |
| 2007 | France | female | 4.621476 | 4.749313 | 4.854717 |
| 2008 | France | female | 4.534260 | 4.658169 | 4.757012 |
| 2009 | France | female | 4.443631 | 4.571115 | 4.668687 |
| 2010 | France | female | 4.352821 | 4.481255 | 4.579245 |
| 2011 | France | female | 4.263869 | 4.393754 | 4.492465 |
| 2012 | France | female | 4.185106 | 4.315402 | 4.412884 |
| 2013 | France | female | 4.108121 | 4.233075 | 4.337252 |
| 2014 | France | female | 4.032322 | 4.155058 | 4.256404 |
| 2015 | France | female | 3.953072 | 4.080264 | 4.177067 |
| 2016 | France | female | 3.874256 | 4.005954 | 4.104518 |
| 2017 | France | female | 3.797011 | 3.929962 | 4.034855 |
| 2018 | France | female | 3.721306 | 3.855256 | 3.968500 |
| 2019 | France | female | 3.647111 | 3.783043 | 3.904226 |
| 2020 | France | female | 3.574809 | 3.709246 | 3.840994 |
| 2021 | France | female | 3.502502 | 3.642894 | 3.778785 |
| 2022 | France | female | 3.431820 | 3.574585 | 3.717584 |
| 2017 | Finland | female | 2.031178 | 2.149249 | 2.250289 |
| 2018 | Finland | female | 2.058736 | 2.175331 | 2.270138 |
| 2019 | Finland | female | 2.073179 | 2.197434 | 2.338795 |
| 2020 | Finland | female | 2.062621 | 2.218542 | 2.405112 |
| 2021 | Finland | female | 2.045617 | 2.246777 | 2.472311 |

*(Continued on Next Page...)*

Table S2: Estimate NHV by country and sex from 2004-2019 (*continued*)
| year | country | sex | 0.025quant | median | 0.975quant |
| --- | --- | --- | --- | --- | --- |
| 2022 | Finland | female | 2.023181 | 2.272947 | 2.547662 |
| 2004 | Greece | female | 3.086101 | 3.226197 | 3.356151 |
| 2005 | Greece | female | 3.066892 | 3.196781 | 3.325205 |
| 2006 | Greece | female | 3.050109 | 3.169539 | 3.294889 |
| 2007 | Greece | female | 3.033393 | 3.142664 | 3.258280 |
| 2008 | Greece | female | 3.016713 | 3.115604 | 3.222211 |
| 2009 | Greece | female | 2.995061 | 3.091100 | 3.190692 |
| 2010 | Greece | female | 2.971392 | 3.066188 | 3.163412 |
| 2011 | Greece | female | 2.944319 | 3.041964 | 3.136675 |
| 2012 | Greece | female | 2.917457 | 3.014951 | 3.110175 |
| 2013 | Greece | female | 2.890841 | 2.988486 | 3.080325 |
| 2014 | Greece | female | 2.864467 | 2.960698 | 3.054306 |
| 2015 | Greece | female | 2.838334 | 2.930976 | 3.028508 |
| 2016 | Greece | female | 2.812440 | 2.905751 | 3.003714 |
| 2017 | Greece | female | 2.786782 | 2.879803 | 2.982835 |
| 2018 | Greece | female | 2.760716 | 2.854590 | 2.957939 |
| 2019 | Greece | female | 2.733438 | 2.829992 | 2.933250 |
| 2020 | Greece | female | 2.706431 | 2.810920 | 2.908768 |
| 2021 | Greece | female | 2.680167 | 2.785614 | 2.885156 |
| 2022 | Greece | female | 2.655134 | 2.763126 | 2.866030 |
| 2015 | Croatia | female | 3.662954 | 3.855509 | 4.056894 |
| 2016 | Croatia | female | 3.780372 | 3.953263 | 4.136959 |
| 2017 | Croatia | female | 3.904693 | 4.054573 | 4.226656 |
| 2018 | Croatia | female | 3.990915 | 4.163482 | 4.357730 |
| 2019 | Croatia | female | 4.079011 | 4.268547 | 4.511060 |
| 2020 | Croatia | female | 4.163259 | 4.399341 | 4.657319 |
| 2021 | Croatia | female | 4.231980 | 4.530046 | 4.826874 |
| 2022 | Croatia | female | 4.283629 | 4.642097 | 5.021880 |
| 2011 | Hungary | female | 4.349776 | 4.571719 | 4.764499 |
| 2012 | Hungary | female | 4.309988 | 4.513142 | 4.683895 |
| 2013 | Hungary | female | 4.271592 | 4.460993 | 4.614039 |
| 2014 | Hungary | female | 4.235215 | 4.411618 | 4.555548 |
| 2015 | Hungary | female | 4.199156 | 4.349337 | 4.494475 |
| 2016 | Hungary | female | 4.129226 | 4.305021 | 4.449953 |
| 2017 | Hungary | female | 4.052095 | 4.249679 | 4.403086 |
| 2018 | Hungary | female | 3.971077 | 4.187805 | 4.364077 |

*(Continued on Next Page...)*

Table S2: Estimate NHV by country and sex from 2004-2019 (*continued*)
| year | country | sex | 0.025quant | median | 0.975quant |
| --- | --- | --- | --- | --- | --- |
| 2019 | Hungary | female | 3.892660 | 4.132993 | 4.335398 |
| 2020 | Hungary | female | 3.818134 | 4.084492 | 4.311446 |
| 2021 | Hungary | female | 3.745035 | 4.039111 | 4.287633 |
| 2022 | Hungary | female | 3.673336 | 3.987933 | 4.263960 |
| 2004 | Italy | female | 5.003464 | 5.146865 | 5.354525 |
| 2005 | Italy | female | 4.969202 | 5.107559 | 5.301872 |
| 2006 | Italy | female | 4.935175 | 5.069477 | 5.249347 |
| 2007 | Italy | female | 4.901381 | 5.030455 | 5.192950 |
| 2008 | Italy | female | 4.867818 | 4.991574 | 5.137161 |
| 2009 | Italy | female | 4.831634 | 4.953935 | 5.083678 |
| 2010 | Italy | female | 4.793832 | 4.915164 | 5.038097 |
| 2011 | Italy | female | 4.756199 | 4.876081 | 4.995010 |
| 2012 | Italy | female | 4.717633 | 4.837252 | 4.952291 |
| 2013 | Italy | female | 4.678586 | 4.802915 | 4.909938 |
| 2014 | Italy | female | 4.639624 | 4.767496 | 4.867947 |
| 2015 | Italy | female | 4.595683 | 4.729644 | 4.826315 |
| 2016 | Italy | female | 4.555452 | 4.692541 | 4.789670 |
| 2017 | Italy | female | 4.515718 | 4.654662 | 4.765324 |
| 2018 | Italy | female | 4.476575 | 4.618528 | 4.740883 |
| 2019 | Italy | female | 4.436312 | 4.583231 | 4.714252 |
| 2020 | Italy | female | 4.395896 | 4.548069 | 4.687772 |
| 2021 | Italy | female | 4.353507 | 4.512817 | 4.661439 |
| 2022 | Italy | female | 4.311373 | 4.474869 | 4.635255 |
| 2017 | Lithuania | female | 2.567954 | 2.702133 | 2.874736 |
| 2018 | Lithuania | female | 2.711482 | 2.852865 | 3.012051 |
| 2019 | Lithuania | female | 2.841251 | 3.007971 | 3.182122 |
| 2020 | Lithuania | female | 2.969932 | 3.174886 | 3.403856 |
| 2021 | Lithuania | female | 3.099896 | 3.347179 | 3.643614 |
| 2022 | Lithuania | female | 3.199185 | 3.501790 | 3.914059 |
| 2013 | Luxembourg | female | 4.726477 | 4.949570 | 5.267130 |
| 2014 | Luxembourg | female | 4.814400 | 5.058309 | 5.355194 |
| 2015 | Luxembourg | female | 4.919486 | 5.158378 | 5.426783 |
| 2016 | Luxembourg | female | 5.007898 | 5.263633 | 5.548727 |
| 2017 | Luxembourg | female | 5.090465 | 5.368788 | 5.646570 |
| 2018 | Luxembourg | female | 5.158050 | 5.484543 | 5.805448 |
| 2019 | Luxembourg | female | 5.202798 | 5.601645 | 5.967219 |

*(Continued on Next Page...)*

Table S2: Estimate NHV by country and sex from 2004-2019 (*continued*)
| year | country | sex | 0.025quant | median | 0.975quant |
| --- | --- | --- | --- | --- | --- |
| 2020 | Luxembourg | female | 5.259403 | 5.713808 | 6.102131 |
| 2021 | Luxembourg | female | 5.316630 | 5.831470 | 6.244278 |
| 2022 | Luxembourg | female | 5.374484 | 5.957734 | 6.409344 |
| 2017 | Latvia | female | 2.535846 | 2.677892 | 2.811642 |
| 2018 | Latvia | female | 2.640800 | 2.792581 | 2.938614 |
| 2019 | Latvia | female | 2.749407 | 2.914481 | 3.056635 |
| 2020 | Latvia | female | 2.834098 | 3.028720 | 3.222079 |
| 2021 | Latvia | female | 2.913740 | 3.168969 | 3.415335 |
| 2022 | Latvia | female | 2.997064 | 3.310926 | 3.632609 |
| 2017 | Malta | female | 2.377570 | 2.555494 | 2.711631 |
| 2018 | Malta | female | 2.376073 | 2.539929 | 2.686867 |
| 2019 | Malta | female | 2.347676 | 2.524545 | 2.679804 |
| 2020 | Malta | female | 2.311834 | 2.513243 | 2.682229 |
| 2021 | Malta | female | 2.256573 | 2.493475 | 2.700241 |
| 2022 | Malta | female | 2.196017 | 2.479374 | 2.728162 |
| 2004 | Netherlands | female | 2.602690 | 2.705632 | 2.824526 |
| 2005 | Netherlands | female | 2.659001 | 2.759838 | 2.874629 |
| 2006 | Netherlands | female | 2.717502 | 2.819180 | 2.922157 |
| 2007 | Netherlands | female | 2.777291 | 2.874020 | 2.973891 |
| 2008 | Netherlands | female | 2.838394 | 2.933740 | 3.038758 |
| 2009 | Netherlands | female | 2.902079 | 2.988639 | 3.098946 |
| 2010 | Netherlands | female | 2.965532 | 3.049264 | 3.159039 |
| 2011 | Netherlands | female | 3.030461 | 3.110214 | 3.218458 |
| 2012 | Netherlands | female | 3.095581 | 3.176008 | 3.282548 |
| 2013 | Netherlands | female | 3.157581 | 3.241965 | 3.348358 |
| 2014 | Netherlands | female | 3.218416 | 3.311928 | 3.424295 |
| 2015 | Netherlands | female | 3.279749 | 3.379642 | 3.497853 |
| 2016 | Netherlands | female | 3.342252 | 3.444922 | 3.573988 |
| 2017 | Netherlands | female | 3.406840 | 3.514502 | 3.653512 |
| 2018 | Netherlands | female | 3.473827 | 3.588371 | 3.738175 |
| 2019 | Netherlands | female | 3.538273 | 3.659044 | 3.825366 |
| 2020 | Netherlands | female | 3.604024 | 3.733569 | 3.923212 |
| 2021 | Netherlands | female | 3.671078 | 3.807553 | 4.023717 |
| 2022 | Netherlands | female | 3.739123 | 3.881499 | 4.126797 |
| 2006 | Poland | female | 4.032279 | 4.232842 | 4.537913 |
| 2007 | Poland | female | 4.022716 | 4.209174 | 4.488761 |

*(Continued on Next Page...)*

Table S2: Estimate NHV by country and sex from 2004-2019 (*continued*)
| year | country | sex | 0.025quant | median | 0.975quant |
| --- | --- | --- | --- | --- | --- |
| 2008 | Poland | female | 4.013176 | 4.187799 | 4.442965 |
| 2009 | Poland | female | 4.001438 | 4.165724 | 4.397641 |
| 2010 | Poland | female | 3.986298 | 4.149503 | 4.352784 |
| 2011 | Poland | female | 3.979180 | 4.126305 | 4.307338 |
| 2012 | Poland | female | 3.974777 | 4.109124 | 4.264534 |
| 2013 | Poland | female | 3.963227 | 4.087104 | 4.235236 |
| 2014 | Poland | female | 3.950580 | 4.067836 | 4.205268 |
| 2015 | Poland | female | 3.937463 | 4.046506 | 4.170202 |
| 2016 | Poland | female | 3.918957 | 4.031999 | 4.138782 |
| 2017 | Poland | female | 3.894375 | 4.006397 | 4.120388 |
| 2018 | Poland | female | 3.866016 | 3.986087 | 4.105609 |
| 2019 | Poland | female | 3.834256 | 3.967192 | 4.084215 |
| 2020 | Poland | female | 3.802758 | 3.942779 | 4.074831 |
| 2021 | Poland | female | 3.769017 | 3.921824 | 4.067606 |
| 2022 | Poland | female | 3.736439 | 3.899084 | 4.062755 |
| 2011 | Portugal | female | 2.853600 | 2.993550 | 3.171013 |
| 2012 | Portugal | female | 2.848492 | 2.970889 | 3.143723 |
| 2013 | Portugal | female | 2.823703 | 2.937229 | 3.115895 |
| 2014 | Portugal | female | 2.791781 | 2.911360 | 3.076875 |
| 2015 | Portugal | female | 2.750132 | 2.896809 | 3.043829 |
| 2016 | Portugal | female | 2.714950 | 2.872076 | 3.026457 |
| 2017 | Portugal | female | 2.680219 | 2.846310 | 3.021533 |
| 2018 | Portugal | female | 2.632801 | 2.817884 | 3.028163 |
| 2019 | Portugal | female | 2.577436 | 2.794656 | 3.016033 |
| 2020 | Portugal | female | 2.523136 | 2.768821 | 3.003956 |
| 2021 | Portugal | female | 2.469982 | 2.746178 | 2.991932 |
| 2022 | Portugal | female | 2.418891 | 2.720391 | 2.979961 |
| 2017 | Romania | female | 2.486490 | 2.638569 | 2.798828 |
| 2018 | Romania | female | 2.631490 | 2.760667 | 2.921779 |
| 2019 | Romania | female | 2.763753 | 2.890966 | 3.056924 |
| 2020 | Romania | female | 2.884204 | 3.023422 | 3.190795 |
| 2021 | Romania | female | 2.981762 | 3.160975 | 3.411926 |
| 2022 | Romania | female | 3.078163 | 3.311118 | 3.627887 |
| 2004 | Sweden | female | 1.743347 | 1.822340 | 1.890626 |
| 2005 | Sweden | female | 1.790677 | 1.864647 | 1.931379 |
| 2006 | Sweden | female | 1.839293 | 1.907955 | 1.973010 |

*(Continued on Next Page...)*

Table S2: Estimate NHV by country and sex from 2004-2019 (*continued*)
| year | country | sex | 0.025quant | median | 0.975quant |
| --- | --- | --- | --- | --- | --- |
| 2007 | Sweden | female | 1.889229 | 1.953785 | 2.015539 |
| 2008 | Sweden | female | 1.940252 | 2.002405 | 2.060917 |
| 2009 | Sweden | female | 1.991951 | 2.053001 | 2.110539 |
| 2010 | Sweden | female | 2.045027 | 2.105433 | 2.159884 |
| 2011 | Sweden | female | 2.099472 | 2.158224 | 2.213027 |
| 2012 | Sweden | female | 2.153431 | 2.210306 | 2.268493 |
| 2013 | Sweden | female | 2.206032 | 2.266084 | 2.325078 |
| 2014 | Sweden | female | 2.260673 | 2.321541 | 2.383097 |
| 2015 | Sweden | female | 2.318576 | 2.378664 | 2.442661 |
| 2016 | Sweden | female | 2.372628 | 2.439669 | 2.508474 |
| 2017 | Sweden | female | 2.427096 | 2.497659 | 2.576503 |
| 2018 | Sweden | female | 2.482761 | 2.556905 | 2.646281 |
| 2019 | Sweden | female | 2.539709 | 2.618921 | 2.721636 |
| 2020 | Sweden | female | 2.594627 | 2.680676 | 2.799340 |
| 2021 | Sweden | female | 2.650392 | 2.744825 | 2.879263 |
| 2022 | Sweden | female | 2.707356 | 2.817257 | 2.961468 |
| 2011 | Slovenia | female | 2.638634 | 2.766283 | 2.851495 |
| 2012 | Slovenia | female | 2.676077 | 2.786193 | 2.865845 |
| 2013 | Slovenia | female | 2.711739 | 2.811347 | 2.880106 |
| 2014 | Slovenia | female | 2.740556 | 2.839933 | 2.905399 |
| 2015 | Slovenia | female | 2.776596 | 2.862361 | 2.934632 |
| 2016 | Slovenia | female | 2.802885 | 2.889001 | 2.963710 |
| 2017 | Slovenia | female | 2.838022 | 2.917714 | 2.996552 |
| 2018 | Slovenia | female | 2.864151 | 2.940017 | 3.035647 |
| 2019 | Slovenia | female | 2.883440 | 2.962796 | 3.078287 |
| 2020 | Slovenia | female | 2.899387 | 2.993771 | 3.115848 |
| 2021 | Slovenia | female | 2.908510 | 3.025173 | 3.156026 |
| 2022 | Slovenia | female | 2.917295 | 3.057086 | 3.199539 |
| 2017 | Slovakia | female | 3.352496 | 3.547030 | 3.760956 |
| 2018 | Slovakia | female | 3.545181 | 3.731166 | 3.944140 |
| 2019 | Slovakia | female | 3.698529 | 3.921240 | 4.131305 |
| 2020 | Slovakia | female | 3.838485 | 4.129527 | 4.428900 |
| 2021 | Slovakia | female | 3.984441 | 4.345077 | 4.734480 |
| 2022 | Slovakia | female | 4.135798 | 4.589135 | 5.085681 |

**Table S3:** Estimate NHV by epidemic period.

| country | sex | period | 0.025quant | median | 0.975quant |
| --- | --- | --- | --- | --- | --- |
| Austria | male | Epidemic | 0.2961026 | 0.3247069 | 0.3652698 |
| Bulgaria | male | Epidemic | 0.1389198 | 0.1589565 | 0.1893942 |
| Belgium | male | Epidemic | 0.2676780 | 0.2938237 | 0.3168746 |
| Cyprus | male | Epidemic | 0.1381502 | 0.1759684 | 0.2235481 |
| Czechia | male | Epidemic | 0.1863108 | 0.2094735 | 0.2302330 |
| Switzerland | male | Epidemic | 0.2514801 | 0.2815730 | 0.3118547 |
| Germany | male | Epidemic | 0.4073919 | 0.4527449 | 0.4930440 |
| Denmark | male | Epidemic | 0.2845631 | 0.3158190 | 0.3570682 |
| Estonia | male | Epidemic | 0.0802252 | 0.0878454 | 0.0978270 |
| Spain | male | Epidemic | 0.1903019 | 0.2093903 | 0.2421676 |
| France | male | Epidemic | 0.4052236 | 0.4399576 | 0.4828990 |
| Finland | male | Epidemic | 0.3531941 | 0.4059161 | 0.4850489 |
| Greece | male | Epidemic | 0.1338503 | 0.1444834 | 0.1596465 |
| Croatia | male | Epidemic | 0.0618355 | 0.0713298 | 0.0836418 |
| Hungary | male | Epidemic | 0.0836844 | 0.0982155 | 0.1232349 |
| Italy | male | Epidemic | 0.0583614 | 0.0631818 | 0.0702774 |
| Lithuania | male | Epidemic | 0.0930986 | 0.1072762 | 0.1250641 |
| Luxembourg | male | Epidemic | 0.1078166 | 0.1251759 | 0.1456070 |
| Latvia | male | Epidemic | 0.1277719 | 0.1528351 | 0.1765361 |
| Malta | male | Epidemic | 0.1543045 | 0.1852309 | 0.2188238 |
| Netherlands | male | Epidemic | 0.1516117 | 0.1737363 | 0.2078641 |
| Poland | male | Epidemic | 0.1424516 | 0.1566970 | 0.1722637 |
| Portugal | male | Epidemic | 0.1830262 | 0.2182611 | 0.2640315 |
| Romania | male | Epidemic | 0.1614400 | 0.1928857 | 0.2182646 |
| Sweden | male | Epidemic | 0.3168408 | 0.3538661 | 0.3993035 |
| Slovenia | male | Epidemic | 0.1150790 | 0.1311057 | 0.1460913 |
| Slovakia | male | Epidemic | 0.0930273 | 0.1086417 | 0.1255407 |
| Austria | male | Post-Epidemic | 0.8530829 | 0.9099770 | 0.9743933 |
| Bulgaria | male | Post-Epidemic | 0.7823400 | 0.9177289 | 1.0831375 |
| Belgium | male | Post-Epidemic | 0.9876400 | 1.0568858 | 1.1170280 |
| Cyprus | male | Post-Epidemic | 1.1772485 | 1.3574490 | 1.5908075 |
| Czechia | male | Post-Epidemic | 1.0526479 | 1.1321710 | 1.2019796 |
| Switzerland | male | Post-Epidemic | 0.9279105 | 1.0115468 | 1.0885385 |
| Germany | male | Post-Epidemic | 1.0606909 | 1.1125397 | 1.1676724 |
| Denmark | male | Post-Epidemic | 0.8544293 | 0.9177864 | 0.9828771 |
| Estonia | male | Post-Epidemic | 1.0618199 | 1.1277465 | 1.2060713 |

*(Continued on Next Page...)*

Table S3: Estimate NHV by epidemic period (*continued*)
| country | sex | period | 0.025quant | median | 0.975quant |
| --- | --- | --- | --- | --- | --- |
| Spain | male | Post-Epidemic | 0.9280577 | 1.0082207 | 1.0901918 |
| France | male | Post-Epidemic | 1.0617575 | 1.1214084 | 1.1929081 |
| Finland | male | Post-Epidemic | 0.8313261 | 0.9560583 | 1.1258789 |
| Greece | male | Post-Epidemic | 0.9942394 | 1.0576204 | 1.1224754 |
| Croatia | male | Post-Epidemic | 0.8458377 | 0.9362992 | 1.0273226 |
| Hungary | male | Post-Epidemic | 0.7822745 | 0.8747332 | 0.9494905 |
| Italy | male | Post-Epidemic | 0.8603274 | 0.9059928 | 0.9544611 |
| Lithuania | male | Post-Epidemic | 0.8358249 | 0.9385477 | 1.0637289 |
| Luxembourg | male | Post-Epidemic | 0.7373418 | 0.8069946 | 0.8905483 |
| Latvia | male | Post-Epidemic | 0.7347193 | 0.8916349 | 1.0361230 |
| Malta | male | Post-Epidemic | 0.7677525 | 0.8893956 | 1.0293716 |
| Netherlands | male | Post-Epidemic | 0.8447200 | 0.9104174 | 0.9732034 |
| Poland | male | Post-Epidemic | 0.7950836 | 0.8511499 | 0.9138764 |
| Portugal | male | Post-Epidemic | 0.8181195 | 0.9463074 | 1.0935674 |
| Romania | male | Post-Epidemic | 0.7791793 | 0.8719217 | 1.0127820 |
| Sweden | male | Post-Epidemic | 0.9060374 | 0.9660835 | 1.0278839 |
| Slovenia | male | Post-Epidemic | 0.8254595 | 0.9041156 | 0.9735542 |
| Slovakia | male | Post-Epidemic | 0.5620056 | 0.6406490 | 0.7110576 |
| Austria | female | Epidemic | 0.2837441 | 0.3068262 | 0.3317178 |
| Bulgaria | female | Epidemic | 0.1291488 | 0.1500241 | 0.1718772 |
| Belgium | female | Epidemic | 0.2207433 | 0.2340601 | 0.2484315 |
| Cyprus | female | Epidemic | 0.0950042 | 0.1135314 | 0.1340445 |
| Czechia | female | Epidemic | 0.1671847 | 0.1791001 | 0.1917894 |
| Switzerland | female | Epidemic | 0.2034044 | 0.2244011 | 0.2410499 |
| Germany | female | Epidemic | 0.3927454 | 0.4196070 | 0.4479282 |
| Denmark | female | Epidemic | 0.2805334 | 0.3055506 | 0.3300202 |
| Estonia | female | Epidemic | 0.0712495 | 0.0782305 | 0.0842722 |
| Spain | female | Epidemic | 0.1682595 | 0.1819127 | 0.2007120 |
| France | female | Epidemic | 0.3525840 | 0.3874634 | 0.4147254 |
| Finland | female | Epidemic | 0.3581683 | 0.4055745 | 0.4508862 |
| Greece | female | Epidemic | 0.1057749 | 0.1136848 | 0.1224595 |
| Croatia | female | Epidemic | 0.0578211 | 0.0651453 | 0.0749464 |
| Hungary | female | Epidemic | 0.0766009 | 0.0888672 | 0.1025201 |
| Italy | female | Epidemic | 0.0470562 | 0.0511942 | 0.0551493 |
| Lithuania | female | Epidemic | 0.0929536 | 0.1048487 | 0.1191917 |
| Luxembourg | female | Epidemic | 0.0964758 | 0.1087169 | 0.1261391 |

*(Continued on Next Page...)*

Table S3: Estimate NHV by epidemic period (*continued*)
| country | sex | period | 0.025quant | median | 0.975quant |
| --- | --- | --- | --- | --- | --- |
| Latvia | female | Epidemic | 0.1065353 | 0.1249193 | 0.1418781 |
| Malta | female | Epidemic | 0.1479663 | 0.1649290 | 0.1891140 |
| Netherlands | female | Epidemic | 0.1403771 | 0.1586452 | 0.1900386 |
| Poland | female | Epidemic | 0.1270995 | 0.1359868 | 0.1483919 |
| Portugal | female | Epidemic | 0.1736436 | 0.1994242 | 0.2315088 |
| Romania | female | Epidemic | 0.1728846 | 0.1927035 | 0.2154959 |
| Sweden | female | Epidemic | 0.2800377 | 0.3115416 | 0.3483092 |
| Slovenia | female | Epidemic | 0.1117062 | 0.1205907 | 0.1305004 |
| Slovakia | female | Epidemic | 0.0989316 | 0.1143543 | 0.1343923 |
| Austria | female | Post-Epidemic | 0.8970601 | 0.9537846 | 0.9995732 |
| Bulgaria | female | Post-Epidemic | 0.8831858 | 0.9816891 | 1.0838686 |
| Belgium | female | Post-Epidemic | 1.0050206 | 1.0457638 | 1.0793023 |
| Cyprus | female | Post-Epidemic | 1.0161588 | 1.1447752 | 1.2968389 |
| Czechia | female | Post-Epidemic | 1.0380854 | 1.0911240 | 1.1511960 |
| Switzerland | female | Post-Epidemic | 0.9392716 | 0.9991395 | 1.0668715 |
| Germany | female | Post-Epidemic | 1.0386183 | 1.0915676 | 1.1482201 |
| Denmark | female | Post-Epidemic | 0.8576652 | 0.9226262 | 0.9829440 |
| Estonia | female | Post-Epidemic | 1.0423757 | 1.0938975 | 1.1456008 |
| Spain | female | Post-Epidemic | 0.9399712 | 1.0020520 | 1.0657079 |
| France | female | Post-Epidemic | 1.0603268 | 1.1067383 | 1.1602217 |
| Finland | female | Post-Epidemic | 0.8455450 | 0.9383145 | 1.0711127 |
| Greece | female | Post-Epidemic | 0.9760275 | 1.0199925 | 1.0700552 |
| Croatia | female | Post-Epidemic | 0.8539702 | 0.9280852 | 1.0061937 |
| Hungary | female | Post-Epidemic | 0.8440750 | 0.9161328 | 0.9977147 |
| Italy | female | Post-Epidemic | 0.8682464 | 0.9125841 | 0.9631327 |
| Lithuania | female | Post-Epidemic | 0.9462452 | 1.0370617 | 1.1307461 |
| Luxembourg | female | Post-Epidemic | 0.7767598 | 0.8491808 | 0.9368589 |
| Latvia | female | Post-Epidemic | 0.8257720 | 0.9113118 | 1.0012395 |
| Malta | female | Post-Epidemic | 0.8062984 | 0.8945664 | 1.0215910 |
| Netherlands | female | Post-Epidemic | 0.8102648 | 0.8620882 | 0.9175611 |
| Poland | female | Post-Epidemic | 0.7668290 | 0.8040323 | 0.8430228 |
| Portugal | female | Post-Epidemic | 0.8756009 | 0.9820132 | 1.0843554 |
| Romania | female | Post-Epidemic | 0.8100965 | 0.8870977 | 0.9617592 |
| Sweden | female | Post-Epidemic | 0.9243669 | 0.9827051 | 1.0419995 |
| Slovenia | female | Post-Epidemic | 0.8927724 | 0.9400193 | 0.9970225 |
| Slovakia | female | Post-Epidemic | 0.6406450 | 0.7149784 | 0.7858796 |

**Table S4:** Estimate relative change by age and epidemic period.

| age | sex | period | 0.025quant | median | 0.975quant |
| --- | --- | --- | --- | --- | --- |
| 50 | male | Pre- | 0.6222053 | 0.6533698 | 0.6833842 |
| 51 | male | Pre- | 0.6416818 | 0.6656251 | 0.6879020 |
| 52 | male | Pre- | 0.6592677 | 0.6815937 | 0.6994560 |
| 53 | male | Pre- | 0.6776096 | 0.6970969 | 0.7142328 |
| 54 | male | Pre- | 0.6924641 | 0.7139882 | 0.7333720 |
| 55 | male | Pre- | 0.7089068 | 0.7311390 | 0.7527415 |
| 56 | male | Pre- | 0.7289271 | 0.7495422 | 0.7696026 |
| 57 | male | Pre- | 0.7505231 | 0.7690813 | 0.7868127 |
| 58 | male | Pre- | 0.7662270 | 0.7854083 | 0.8074882 |
| 59 | male | Pre- | 0.7804060 | 0.8007088 | 0.8222629 |
| 60 | male | Pre- | 0.7956348 | 0.8142463 | 0.8348948 |
| 61 | male | Pre- | 0.8042011 | 0.8257226 | 0.8456175 |
| 62 | male | Pre- | 0.8157176 | 0.8369881 | 0.8554958 |
| 63 | male | Pre- | 0.8301705 | 0.8472905 | 0.8687556 |
| 64 | male | Pre- | 0.8416070 | 0.8598079 | 0.8795001 |
| 65 | male | Pre- | 0.8542529 | 0.8732766 | 0.8938377 |
| 66 | male | Pre- | 0.8677460 | 0.8894433 | 0.9065529 |
| 67 | male | Pre- | 0.8849658 | 0.9048353 | 0.9252388 |
| 68 | male | Pre- | 0.9047495 | 0.9224522 | 0.9430781 |
| 69 | male | Pre- | 0.9158714 | 0.9405733 | 0.9618433 |
| 70 | male | Pre- | 0.9318820 | 0.9566880 | 0.9795191 |
| 71 | male | Pre- | 0.9485974 | 0.9737198 | 0.9954537 |
| 72 | male | Pre- | 0.9658392 | 0.9894204 | 1.0105642 |
| 73 | male | Pre- | 0.9835678 | 1.0058701 | 1.0296385 |
| 74 | male | Pre- | 0.9970748 | 1.0248482 | 1.0449809 |
| 75 | male | Pre- | 1.0137271 | 1.0409441 | 1.0617639 |
| 76 | male | Pre- | 1.0308115 | 1.0581002 | 1.0811283 |
| 77 | male | Pre- | 1.0453467 | 1.0746535 | 1.0995508 |
| 78 | male | Pre- | 1.0629160 | 1.0924531 | 1.1202138 |
| 79 | male | Pre- | 1.0785049 | 1.1101039 | 1.1348833 |
| 80 | male | Pre- | 1.0944649 | 1.1267834 | 1.1555908 |
| 81 | male | Pre- | 1.1049074 | 1.1420933 | 1.1727871 |
| 82 | male | Pre- | 1.1198229 | 1.1594414 | 1.1901931 |
| 83 | male | Pre- | 1.1361074 | 1.1732528 | 1.2098521 |
| 84 | male | Pre- | 1.1477130 | 1.1849660 | 1.2216739 |
| 85 | male | Pre- | 1.1579440 | 1.1976237 | 1.2324224 |

*(Continued on Next Page...)*

Table S4: Estimate relative change by age and epidemic period (*continued*)
| age | sex | period | 0.025quant | median | 0.975quant |
| --- | --- | --- | --- | --- | --- |
| 86 | male | Pre- | 1.1734826 | 1.2093617 | 1.2386285 |
| 87 | male | Pre- | 1.1816384 | 1.2175757 | 1.2472776 |
| 88 | male | Pre- | 1.1866461 | 1.2235984 | 1.2584300 |
| 89 | male | Pre- | 1.1899392 | 1.2303481 | 1.2616889 |
| 90 | male | Pre- | 1.1946845 | 1.2375402 | 1.2724510 |
| 91 | male | Pre- | 1.2013122 | 1.2416181 | 1.2810022 |
| 92 | male | Pre- | 1.2100312 | 1.2459908 | 1.2920233 |
| 93 | male | Pre- | 1.2128703 | 1.2542417 | 1.3035071 |
| 94 | male | Pre- | 1.2133847 | 1.2625152 | 1.3279619 |
| 95 | male | Pre- | 1.2099136 | 1.2688088 | 1.3506962 |
| 96 | male | Pre- | 1.1999127 | 1.2703347 | 1.3713414 |
| 97 | male | Pre- | 1.1931920 | 1.2741572 | 1.3981606 |
| 98 | male | Pre- | 1.1761062 | 1.2811552 | 1.4326137 |
| 99 | male | Pre- | 1.1560347 | 1.2911998 | 1.4580449 |
| 100 | male | Pre- | 1.1344307 | 1.3053740 | 1.4773485 |
| 50 | male | Epi- | 0.6883154 | 0.8104052 | 0.9898002 |
| 51 | male | Epi- | 0.7142334 | 0.8246640 | 0.9841284 |
| 52 | male | Epi- | 0.7357728 | 0.8389158 | 0.9748656 |
| 53 | male | Epi- | 0.7622906 | 0.8501604 | 0.9591878 |
| 54 | male | Epi- | 0.7896488 | 0.8654134 | 0.9486859 |
| 55 | male | Epi- | 0.8145691 | 0.8843014 | 0.9509256 |
| 56 | male | Epi- | 0.8380623 | 0.9020646 | 0.9620791 |
| 57 | male | Epi- | 0.8674671 | 0.9172548 | 0.9660522 |
| 58 | male | Epi- | 0.8892424 | 0.9325655 | 0.9830726 |
| 59 | male | Epi- | 0.9065305 | 0.9525965 | 1.0009867 |
| 60 | male | Epi- | 0.9242783 | 0.9694613 | 1.0160338 |
| 61 | male | Epi- | 0.9346854 | 0.9875483 | 1.0322758 |
| 62 | male | Epi- | 0.9463376 | 1.0051316 | 1.0510993 |
| 63 | male | Epi- | 0.9585794 | 1.0207241 | 1.0704292 |
| 64 | male | Epi- | 0.9753638 | 1.0369576 | 1.0930384 |
| 65 | male | Epi- | 0.9924421 | 1.0517322 | 1.1075837 |
| 66 | male | Epi- | 1.0116853 | 1.0665473 | 1.1312740 |
| 67 | male | Epi- | 1.0229519 | 1.0810994 | 1.1497987 |
| 68 | male | Epi- | 1.0336813 | 1.0919364 | 1.1619851 |
| 69 | male | Epi- | 1.0463344 | 1.1022848 | 1.1646383 |
| 70 | male | Epi- | 1.0569905 | 1.1099301 | 1.1764088 |

*(Continued on Next Page...)*

Table S4: Estimate relative change by age and epidemic period (*continued*)
| age | sex | period | 0.025quant | median | 0.975quant |
| --- | --- | --- | --- | --- | --- |
| 71 | male | Epi- | 1.0654733 | 1.1171033 | 1.1892558 |
| 72 | male | Epi- | 1.0717503 | 1.1248830 | 1.1998850 |
| 73 | male | Epi- | 1.0702287 | 1.1314607 | 1.2032970 |
| 74 | male | Epi- | 1.0762820 | 1.1352214 | 1.2055469 |
| 75 | male | Epi- | 1.0869140 | 1.1417025 | 1.2181176 |
| 76 | male | Epi- | 1.0951474 | 1.1472433 | 1.2177877 |
| 77 | male | Epi- | 1.0992321 | 1.1513562 | 1.2281963 |
| 78 | male | Epi- | 1.0971388 | 1.1518003 | 1.2294282 |
| 79 | male | Epi- | 1.0918210 | 1.1510440 | 1.2314422 |
| 80 | male | Epi- | 1.0884366 | 1.1517090 | 1.2267811 |
| 81 | male | Epi- | 1.0833947 | 1.1439691 | 1.2191653 |
| 82 | male | Epi- | 1.0752461 | 1.1365337 | 1.2194371 |
| 83 | male | Epi- | 1.0640121 | 1.1268748 | 1.2063660 |
| 84 | male | Epi- | 1.0550471 | 1.1144722 | 1.1827760 |
| 85 | male | Epi- | 1.0377403 | 1.0991675 | 1.1618227 |
| 86 | male | Epi- | 1.0266644 | 1.0825446 | 1.1346798 |
| 87 | male | Epi- | 1.0119757 | 1.0635615 | 1.1119161 |
| 88 | male | Epi- | 0.9989992 | 1.0495251 | 1.0996934 |
| 89 | male | Epi- | 0.9796118 | 1.0283914 | 1.0818339 |
| 90 | male | Epi- | 0.9489894 | 1.0066059 | 1.0648148 |
| 91 | male | Epi- | 0.9225675 | 0.9834939 | 1.0422294 |
| 92 | male | Epi- | 0.8895485 | 0.9597697 | 1.0192369 |
| 93 | male | Epi- | 0.8545814 | 0.9336864 | 1.0079216 |
| 94 | male | Epi- | 0.8211315 | 0.9108288 | 1.0021571 |
| 95 | male | Epi- | 0.7910191 | 0.8905596 | 0.9956785 |
| 96 | male | Epi- | 0.7566473 | 0.8662451 | 0.9879954 |
| 97 | male | Epi- | 0.7281744 | 0.8513442 | 0.9848695 |
| 98 | male | Epi- | 0.6910998 | 0.8299928 | 0.9813572 |
| 99 | male | Epi- | 0.6523472 | 0.8123359 | 0.9822552 |
| 100 | male | Epi- | 0.6160206 | 0.7910277 | 0.9797077 |
| 50 | male | Post- | 0.6707334 | 0.7630509 | 0.8367007 |
| 51 | male | Post- | 0.6926570 | 0.7641664 | 0.8256209 |
| 52 | male | Post- | 0.7057793 | 0.7695621 | 0.8164827 |
| 53 | male | Post- | 0.7246961 | 0.7726911 | 0.8187910 |
| 54 | male | Post- | 0.7400303 | 0.7794174 | 0.8226015 |
| 55 | male | Post- | 0.7527089 | 0.7877303 | 0.8212115 |

*(Continued on Next Page...)*

Table S4: Estimate relative change by age and epidemic period (*continued*)
| age | sex | period | 0.025quant | median | 0.975quant |
| --- | --- | --- | --- | --- | --- |
| 56 | male | Post- | 0.7694026 | 0.7966090 | 0.8244880 |
| 57 | male | Post- | 0.7762740 | 0.8064949 | 0.8333265 |
| 58 | male | Post- | 0.7841614 | 0.8159495 | 0.8442348 |
| 59 | male | Post- | 0.7931043 | 0.8252855 | 0.8546448 |
| 60 | male | Post- | 0.8041475 | 0.8341772 | 0.8653198 |
| 61 | male | Post- | 0.8102126 | 0.8433129 | 0.8752219 |
| 62 | male | Post- | 0.8171678 | 0.8530909 | 0.8846452 |
| 63 | male | Post- | 0.8276646 | 0.8613219 | 0.8936457 |
| 64 | male | Post- | 0.8373883 | 0.8705070 | 0.8988273 |
| 65 | male | Post- | 0.8499301 | 0.8793692 | 0.9089531 |
| 66 | male | Post- | 0.8622299 | 0.8898914 | 0.9213583 |
| 67 | male | Post- | 0.8787459 | 0.9044013 | 0.9364049 |
| 68 | male | Post- | 0.8931209 | 0.9201839 | 0.9511252 |
| 69 | male | Post- | 0.9089324 | 0.9390879 | 0.9734907 |
| 70 | male | Post- | 0.9212508 | 0.9589640 | 0.9963772 |
| 71 | male | Post- | 0.9394459 | 0.9791683 | 1.0155799 |
| 72 | male | Post- | 0.9634720 | 0.9973786 | 1.0322316 |
| 73 | male | Post- | 0.9851485 | 1.0162120 | 1.0503125 |
| 74 | male | Post- | 1.0047512 | 1.0353312 | 1.0714133 |
| 75 | male | Post- | 1.0186554 | 1.0564956 | 1.0842223 |
| 76 | male | Post- | 1.0339273 | 1.0701875 | 1.1015622 |
| 77 | male | Post- | 1.0505662 | 1.0865943 | 1.1212474 |
| 78 | male | Post- | 1.0677691 | 1.0995526 | 1.1319463 |
| 79 | male | Post- | 1.0778076 | 1.1108659 | 1.1399659 |
| 80 | male | Post- | 1.0849875 | 1.1174650 | 1.1527027 |
| 81 | male | Post- | 1.0932975 | 1.1258090 | 1.1652680 |
| 82 | male | Post- | 1.0918235 | 1.1332573 | 1.1713685 |
| 83 | male | Post- | 1.0937156 | 1.1379632 | 1.1747245 |
| 84 | male | Post- | 1.0889706 | 1.1433632 | 1.1839858 |
| 85 | male | Post- | 1.0898756 | 1.1481706 | 1.1814520 |
| 86 | male | Post- | 1.0951679 | 1.1482615 | 1.1906921 |
| 87 | male | Post- | 1.1032524 | 1.1513952 | 1.1993125 |
| 88 | male | Post- | 1.1056941 | 1.1532422 | 1.1967563 |
| 89 | male | Post- | 1.1183448 | 1.1553507 | 1.2033343 |
| 90 | male | Post- | 1.1204613 | 1.1612646 | 1.2077006 |
| 91 | male | Post- | 1.1163682 | 1.1679364 | 1.2144322 |

*(Continued on Next Page...)*

Table S4: Estimate relative change by age and epidemic period (*continued*)
| age | sex | period | 0.025quant | median | 0.975quant |
| --- | --- | --- | --- | --- | --- |
| 92 | male | Post- | 1.1233311 | 1.1712027 | 1.2264122 |
| 93 | male | Post- | 1.1238953 | 1.1775160 | 1.2325316 |
| 94 | male | Post- | 1.1190689 | 1.1833409 | 1.2571602 |
| 95 | male | Post- | 1.1073479 | 1.1950633 | 1.2925261 |
| 96 | male | Post- | 1.1065562 | 1.2082019 | 1.3304914 |
| 97 | male | Post- | 1.0981154 | 1.2135210 | 1.3617853 |
| 98 | male | Post- | 1.0982014 | 1.2198747 | 1.4066325 |
| 99 | male | Post- | 1.0856505 | 1.2299197 | 1.4652797 |
| 100 | male | Post- | 1.0695956 | 1.2502039 | 1.5261327 |
| 50 | female | Pre- | 0.7815025 | 0.8119882 | 0.8316218 |
| 51 | female | Pre- | 0.8071039 | 0.8261550 | 0.8452974 |
| 52 | female | Pre- | 0.8243696 | 0.8405848 | 0.8590363 |
| 53 | female | Pre- | 0.8383646 | 0.8547825 | 0.8725730 |
| 54 | female | Pre- | 0.8496527 | 0.8650251 | 0.8830974 |
| 55 | female | Pre- | 0.8573498 | 0.8727603 | 0.8910158 |
| 56 | female | Pre- | 0.8642901 | 0.8786580 | 0.8956651 |
| 57 | female | Pre- | 0.8702012 | 0.8843515 | 0.8988677 |
| 58 | female | Pre- | 0.8736729 | 0.8888036 | 0.9033350 |
| 59 | female | Pre- | 0.8778217 | 0.8933030 | 0.9095484 |
| 60 | female | Pre- | 0.8848391 | 0.8966094 | 0.9119570 |
| 61 | female | Pre- | 0.8872541 | 0.8988580 | 0.9156757 |
| 62 | female | Pre- | 0.8918710 | 0.9038785 | 0.9201624 |
| 63 | female | Pre- | 0.8967339 | 0.9117880 | 0.9263951 |
| 64 | female | Pre- | 0.9006645 | 0.9180494 | 0.9360093 |
| 65 | female | Pre- | 0.9115157 | 0.9284884 | 0.9450184 |
| 66 | female | Pre- | 0.9232702 | 0.9391268 | 0.9539645 |
| 67 | female | Pre- | 0.9343386 | 0.9493078 | 0.9658324 |
| 68 | female | Pre- | 0.9466225 | 0.9598666 | 0.9768599 |
| 69 | female | Pre- | 0.9599895 | 0.9725636 | 0.9910834 |
| 70 | female | Pre- | 0.9732785 | 0.9864411 | 1.0054248 |
| 71 | female | Pre- | 0.9839807 | 1.0007139 | 1.0193345 |
| 72 | female | Pre- | 0.9971176 | 1.0151275 | 1.0341489 |
| 73 | female | Pre- | 1.0113780 | 1.0297433 | 1.0475144 |
| 74 | female | Pre- | 1.0216369 | 1.0419215 | 1.0583797 |
| 75 | female | Pre- | 1.0338095 | 1.0503043 | 1.0690208 |
| 76 | female | Pre- | 1.0423978 | 1.0593416 | 1.0828863 |

*(Continued on Next Page...)*

Table S4: Estimate relative change by age and epidemic period (*continued*)
| age | sex | period | 0.025quant | median | 0.975quant |
| --- | --- | --- | --- | --- | --- |
| 77 | female | Pre- | 1.0550548 | 1.0721081 | 1.0931343 |
| 78 | female | Pre- | 1.0629836 | 1.0798352 | 1.1013940 |
| 79 | female | Pre- | 1.0701044 | 1.0872005 | 1.1107445 |
| 80 | female | Pre- | 1.0759576 | 1.0949800 | 1.1196235 |
| 81 | female | Pre- | 1.0809602 | 1.1015132 | 1.1283404 |
| 82 | female | Pre- | 1.0863399 | 1.1076949 | 1.1310186 |
| 83 | female | Pre- | 1.0887337 | 1.1100642 | 1.1292607 |
| 84 | female | Pre- | 1.0834601 | 1.1041591 | 1.1254115 |
| 85 | female | Pre- | 1.0750049 | 1.0992125 | 1.1225347 |
| 86 | female | Pre- | 1.0646421 | 1.0926805 | 1.1197021 |
| 87 | female | Pre- | 1.0678127 | 1.0850073 | 1.1139623 |
| 88 | female | Pre- | 1.0571061 | 1.0840884 | 1.1142019 |
| 89 | female | Pre- | 1.0552028 | 1.0846351 | 1.1160246 |
| 90 | female | Pre- | 1.0594275 | 1.0859167 | 1.1127814 |
| 91 | female | Pre- | 1.0614107 | 1.0890784 | 1.1149971 |
| 92 | female | Pre- | 1.0540312 | 1.0917476 | 1.1144254 |
| 93 | female | Pre- | 1.0496824 | 1.0903285 | 1.1205498 |
| 94 | female | Pre- | 1.0453982 | 1.0905359 | 1.1295095 |
| 95 | female | Pre- | 1.0389798 | 1.0900603 | 1.1392160 |
| 96 | female | Pre- | 1.0351422 | 1.0895820 | 1.1490104 |
| 97 | female | Pre- | 1.0054758 | 1.0883097 | 1.1553209 |
| 98 | female | Pre- | 0.9873885 | 1.0854842 | 1.1639615 |
| 99 | female | Pre- | 0.9540851 | 1.0856129 | 1.1827220 |
| 100 | female | Pre- | 0.9254002 | 1.0769370 | 1.2069627 |
| 50 | female | Epi- | 0.8706597 | 0.9865528 | 1.1377625 |
| 51 | female | Epi- | 0.9103717 | 1.0079176 | 1.1309209 |
| 52 | female | Epi- | 0.9374478 | 1.0193744 | 1.1285081 |
| 53 | female | Epi- | 0.9657908 | 1.0372702 | 1.1249016 |
| 54 | female | Epi- | 0.9968602 | 1.0535128 | 1.1181576 |
| 55 | female | Epi- | 1.0172019 | 1.0706959 | 1.1240805 |
| 56 | female | Epi- | 1.0422858 | 1.0882191 | 1.1361629 |
| 57 | female | Epi- | 1.0590987 | 1.1033540 | 1.1588060 |
| 58 | female | Epi- | 1.0696543 | 1.1158277 | 1.1677368 |
| 59 | female | Epi- | 1.0866250 | 1.1326183 | 1.1731917 |
| 60 | female | Epi- | 1.0950046 | 1.1436739 | 1.1897839 |
| 61 | female | Epi- | 1.1079754 | 1.1597323 | 1.2046173 |

*(Continued on Next Page...)*

Table S4: Estimate relative change by age and epidemic period (*continued*)
| age | sex | period | 0.025quant | median | 0.975quant |
| --- | --- | --- | --- | --- | --- |
| 62 | female | Epi- | 1.1217777 | 1.1693217 | 1.2140973 |
| 63 | female | Epi- | 1.1339007 | 1.1812069 | 1.2195257 |
| 64 | female | Epi- | 1.1517698 | 1.1968029 | 1.2374915 |
| 65 | female | Epi- | 1.1670144 | 1.2099839 | 1.2557815 |
| 66 | female | Epi- | 1.1774020 | 1.2214487 | 1.2667037 |
| 67 | female | Epi- | 1.1820983 | 1.2327550 | 1.2772745 |
| 68 | female | Epi- | 1.1937011 | 1.2408478 | 1.2839444 |
| 69 | female | Epi- | 1.1984540 | 1.2454778 | 1.2864408 |
| 70 | female | Epi- | 1.2025519 | 1.2459911 | 1.2935315 |
| 71 | female | Epi- | 1.1979456 | 1.2450143 | 1.2971380 |
| 72 | female | Epi- | 1.1981221 | 1.2481858 | 1.3038860 |
| 73 | female | Epi- | 1.1955629 | 1.2445500 | 1.2987913 |
| 74 | female | Epi- | 1.1908260 | 1.2418051 | 1.2986831 |
| 75 | female | Epi- | 1.1890762 | 1.2356508 | 1.2897444 |
| 76 | female | Epi- | 1.1807917 | 1.2289042 | 1.2845050 |
| 77 | female | Epi- | 1.1660568 | 1.2171169 | 1.2719802 |
| 78 | female | Epi- | 1.1573004 | 1.2054077 | 1.2609875 |
| 79 | female | Epi- | 1.1438565 | 1.1888438 | 1.2368196 |
| 80 | female | Epi- | 1.1229975 | 1.1696107 | 1.2140155 |
| 81 | female | Epi- | 1.0986725 | 1.1465074 | 1.1874915 |
| 82 | female | Epi- | 1.0609282 | 1.1213499 | 1.1581826 |
| 83 | female | Epi- | 1.0283878 | 1.0937087 | 1.1326144 |
| 84 | female | Epi- | 0.9994300 | 1.0614431 | 1.1033608 |
| 85 | female | Epi- | 0.9658781 | 1.0222269 | 1.0667208 |
| 86 | female | Epi- | 0.9340138 | 0.9867882 | 1.0338491 |
| 87 | female | Epi- | 0.8990711 | 0.9509264 | 0.9967888 |
| 88 | female | Epi- | 0.8623388 | 0.9121330 | 0.9560862 |
| 89 | female | Epi- | 0.8251271 | 0.8690286 | 0.9169674 |
| 90 | female | Epi- | 0.7876609 | 0.8311484 | 0.8718611 |
| 91 | female | Epi- | 0.7564707 | 0.7909154 | 0.8314868 |
| 92 | female | Epi- | 0.7217907 | 0.7537304 | 0.8009679 |
| 93 | female | Epi- | 0.6728842 | 0.7181364 | 0.7667129 |
| 94 | female | Epi- | 0.6358407 | 0.6826216 | 0.7438450 |
| 95 | female | Epi- | 0.5874116 | 0.6518856 | 0.7214812 |
| 96 | female | Epi- | 0.5443218 | 0.6184683 | 0.7014513 |
| 97 | female | Epi- | 0.5028856 | 0.5888582 | 0.6814435 |

*(Continued on Next Page...)*

Table S4: Estimate relative change by age and epidemic period (*continued*)
| age | sex | period | 0.025quant | median | 0.975quant |
| --- | --- | --- | --- | --- | --- |
| 98 | female | Epi- | 0.4638019 | 0.5588165 | 0.6674853 |
| 99 | female | Epi- | 0.4252971 | 0.5331682 | 0.6556862 |
| 100 | female | Epi- | 0.3914726 | 0.5131565 | 0.6466017 |
| 50 | female | Post- | 0.8561584 | 0.9196215 | 1.0056997 |
| 51 | female | Post- | 0.8696308 | 0.9157466 | 0.9794793 |
| 52 | female | Post- | 0.8768128 | 0.9169513 | 0.9599675 |
| 53 | female | Post- | 0.8837364 | 0.9179982 | 0.9619072 |
| 54 | female | Post- | 0.8900209 | 0.9170189 | 0.9605811 |
| 55 | female | Post- | 0.8933345 | 0.9191002 | 0.9528291 |
| 56 | female | Post- | 0.8999044 | 0.9243166 | 0.9545170 |
| 57 | female | Post- | 0.9056269 | 0.9267017 | 0.9537038 |
| 58 | female | Post- | 0.9067890 | 0.9301585 | 0.9574895 |
| 59 | female | Post- | 0.9099090 | 0.9325629 | 0.9603931 |
| 60 | female | Post- | 0.9103039 | 0.9340127 | 0.9650191 |
| 61 | female | Post- | 0.9055331 | 0.9313394 | 0.9597812 |
| 62 | female | Post- | 0.9014901 | 0.9257688 | 0.9584252 |
| 63 | female | Post- | 0.9007951 | 0.9246952 | 0.9600540 |
| 64 | female | Post- | 0.9065868 | 0.9287671 | 0.9600954 |
| 65 | female | Post- | 0.9144361 | 0.9357648 | 0.9633542 |
| 66 | female | Post- | 0.9240375 | 0.9425745 | 0.9707616 |
| 67 | female | Post- | 0.9330892 | 0.9524920 | 0.9788020 |
| 68 | female | Post- | 0.9449240 | 0.9649880 | 0.9896051 |
| 69 | female | Post- | 0.9564154 | 0.9773756 | 1.0043973 |
| 70 | female | Post- | 0.9696848 | 0.9891217 | 1.0194564 |
| 71 | female | Post- | 0.9771799 | 0.9984705 | 1.0288565 |
| 72 | female | Post- | 0.9883538 | 1.0094447 | 1.0396294 |
| 73 | female | Post- | 0.9941772 | 1.0164871 | 1.0465611 |
| 74 | female | Post- | 0.9975955 | 1.0214106 | 1.0508974 |
| 75 | female | Post- | 1.0046047 | 1.0268858 | 1.0513244 |
| 76 | female | Post- | 1.0054259 | 1.0302623 | 1.0585851 |
| 77 | female | Post- | 1.0054525 | 1.0355958 | 1.0642291 |
| 78 | female | Post- | 1.0086401 | 1.0388097 | 1.0640639 |
| 79 | female | Post- | 1.0140248 | 1.0417650 | 1.0653672 |
| 80 | female | Post- | 1.0181289 | 1.0462843 | 1.0674445 |
| 81 | female | Post- | 1.0208740 | 1.0492659 | 1.0715527 |
| 82 | female | Post- | 1.0234667 | 1.0492613 | 1.0751581 |

*(Continued on Next Page...)*

Table S4: Estimate relative change by age and epidemic period (*continued*)
| age | sex | period | 0.025quant | median | 0.975quant |
| --- | --- | --- | --- | --- | --- |
| 83 | female | Post- | 1.0279543 | 1.0518600 | 1.0793775 |
| 84 | female | Post- | 1.0277758 | 1.0494467 | 1.0796764 |
| 85 | female | Post- | 1.0202654 | 1.0496329 | 1.0780220 |
| 86 | female | Post- | 1.0168089 | 1.0478910 | 1.0753413 |
| 87 | female | Post- | 1.0110457 | 1.0421603 | 1.0756095 |
| 88 | female | Post- | 1.0045175 | 1.0395665 | 1.0735003 |
| 89 | female | Post- | 1.0019788 | 1.0377520 | 1.0804643 |
| 90 | female | Post- | 0.9963083 | 1.0387614 | 1.0845054 |
| 91 | female | Post- | 0.9978090 | 1.0405899 | 1.0900797 |
| 92 | female | Post- | 1.0031960 | 1.0469077 | 1.1002063 |
| 93 | female | Post- | 1.0104450 | 1.0569612 | 1.1098110 |
| 94 | female | Post- | 1.0183289 | 1.0652083 | 1.1371837 |
| 95 | female | Post- | 1.0122996 | 1.0725546 | 1.1579867 |
| 96 | female | Post- | 1.0089082 | 1.0780600 | 1.1748477 |
| 97 | female | Post- | 0.9980404 | 1.0853256 | 1.1877694 |
| 98 | female | Post- | 0.9885380 | 1.0855871 | 1.2118314 |
| 99 | female | Post- | 0.9626645 | 1.0915711 | 1.2313302 |
| 100 | female | Post- | 0.9308025 | 1.0983964 | 1.2661772 |

**Table S5:** Estimate relative change by diseases and epidemic period.

| disease | sex | period | 0.025quant | median | 0.975quant |
| --- | --- | --- | --- | --- | --- |
| Heart attack | male | Pre- | 1.4587876 | 1.4813909 | 1.4988766 |
| Heart attack | male | Epi- | 1.1348368 | 1.1406969 | 1.1451895 |
| Heart attack | male | Post- | 1.2910142 | 1.3045119 | 1.3149078 |
| Hypertension | male | Pre- | 1.3289685 | 1.3441546 | 1.3604443 |
| Hypertension | male | Epi- | 1.1667703 | 1.1739821 | 1.1816768 |
| Hypertension | male | Post- | 1.2754537 | 1.2879117 | 1.3012526 |
| High cholesterol | male | Pre- | 1.0890480 | 1.1023475 | 1.1150636 |
| High cholesterol | male | Epi- | 1.0406806 | 1.0466022 | 1.0522285 |
| High cholesterol | male | Post- | 1.0723555 | 1.0830681 | 1.0932890 |
| Stroke | male | Pre- | 1.3269659 | 1.3482710 | 1.3739982 |
| Stroke | male | Epi- | 1.0055052 | 1.0058160 | 1.0061850 |
| Stroke | male | Post- | 1.2872210 | 1.3056506 | 1.3278639 |
| Diabetes | male | Pre- | 1.4113148 | 1.4311858 | 1.4567106 |

*(Continued on Next Page...)*

Table S5: Estimate relative change by diseases and epidemic period (*continued*)
| disease | sex | period | 0.025quant | median | 0.975quant |
| --- | --- | --- | --- | --- | --- |
| Diabetes | male | Epi- | 1.1310851 | 1.1367534 | 1.1439606 |
| Diabetes | male | Post- | 1.2930419 | 1.3065984 | 1.3239420 |
| Chronic lung disease | male | Pre- | 1.3853012 | 1.4160244 | 1.4441781 |
| Chronic lung disease | male | Epi- | 1.0221289 | 1.0236357 | 1.0249900 |
| Chronic lung disease | male | Post- | 1.2716534 | 1.2923890 | 1.3112867 |
| Cancer | male | Pre- | 1.7504840 | 1.7790278 | 1.8104277 |
| Cancer | male | Epi- | 1.3190351 | 1.3296289 | 1.3411839 |
| Cancer | male | Post- | 1.8702604 | 1.9043950 | 1.9420201 |
| Stomach, duodenal, peptic ulcer | male | Pre- | 1.1589364 | 1.1860163 | 1.2118429 |
| Stomach, duodenal, peptic ulcer | male | Epi- | 1.0923415 | 1.1075541 | 1.1219333 |
| Stomach, duodenal, peptic ulcer | male | Post- | 1.2366789 | 1.2785079 | 1.3187946 |
| Parkinson | male | Pre- | 1.0159426 | 1.0774327 | 1.1256920 |
| Parkinson | male | Epi- | 1.0145814 | 1.0706431 | 1.1144515 |
| Parkinson | male | Post- | 1.0144588 | 1.0700334 | 1.1134441 |
| Cataracts | male | Pre- | 0.9906488 | 1.0264563 | 1.0780816 |
| Cataracts | male | Epi- | 0.9992595 | 1.0020610 | 1.0059455 |
| Cataracts | male | Post- | 0.9830489 | 1.0486636 | 1.1466133 |
| Hip, femoral fracture | male | Pre- | 1.2151848 | 1.2541101 | 1.3073349 |
| Hip, femoral fracture | male | Epi- | 1.0418899 | 1.0488299 | 1.0580485 |
| Hip, femoral fracture | male | Post- | 1.2825910 | 1.3352866 | 1.4080768 |
| Heart attack | female | Pre- | 1.3225599 | 1.3400570 | 1.3579099 |
| Heart attack | female | Epi- | 1.0176509 | 1.0184883 | 1.0193323 |
| Heart attack | female | Post- | 1.2811672 | 1.2961777 | 1.3114703 |
| Hypertension | female | Pre- | 1.2846734 | 1.2944496 | 1.3073456 |
| Hypertension | female | Epi- | 1.0798183 | 1.0823307 | 1.0856248 |
| Hypertension | female | Post- | 1.2288441 | 1.2365316 | 1.2466568 |
| High cholesterol | female | Pre- | 1.1060872 | 1.1186127 | 1.1286852 |
| High cholesterol | female | Epi- | 1.1312700 | 1.1469607 | 1.1596071 |
| High cholesterol | female | Post- | 1.0857977 | 1.0958253 | 1.1038743 |
| Stroke | female | Pre- | 1.2646175 | 1.2953448 | 1.3176041 |
| Stroke | female | Epi- | 1.0388625 | 1.0429208 | 1.0458105 |
| Stroke | female | Post- | 1.1868877 | 1.2078659 | 1.2229787 |
| Diabetes | female | Pre- | 1.3230686 | 1.3420185 | 1.3622144 |
| Diabetes | female | Epi- | 1.1231901 | 1.1298381 | 1.1368629 |
| Diabetes | female | Post- | 1.2203246 | 1.2327305 | 1.2458965 |
| Chronic lung disease | female | Pre- | 1.2968971 | 1.3176336 | 1.3386418 |

*(Continued on Next Page...)*

Table S5: Estimate relative change by diseases and epidemic period (*continued*)
| disease | sex | period | 0.025quant | median | 0.975quant |
| --- | --- | --- | --- | --- | --- |
| Chronic lung disease | female | Epi- | 1.0728648 | 1.0774790 | 1.0820998 |
| Chronic lung disease | female | Post- | 1.2506895 | 1.2678775 | 1.2852522 |
| Cancer | female | Pre- | 1.5881518 | 1.6170753 | 1.6384253 |
| Cancer | female | Epi- | 1.1328284 | 1.1383544 | 1.1423873 |
| Cancer | female | Post- | 1.6654166 | 1.6988930 | 1.7236435 |
| Stomach, duodenal, peptic ulcer | female | Pre- | 1.1814648 | 1.2064351 | 1.2258661 |
| Stomach, duodenal, peptic ulcer | female | Epi- | 1.0229706 | 1.0258887 | 1.0281235 |
| Stomach, duodenal, peptic ulcer | female | Post- | 1.1947126 | 1.2216702 | 1.2426735 |
| Parkinson | female | Pre- | 0.5195846 | 0.6686785 | 0.8667815 |
| Parkinson | female | Epi- | 0.6524818 | 0.7691662 | 0.9109753 |
| Parkinson | female | Post- | 0.4636418 | 0.6234533 | 0.8454892 |
| Cataracts | female | Pre- | 1.2825368 | 1.6745456 | 2.1614218 |
| Cataracts | female | Epi- | 1.1208436 | 1.2666501 | 1.4238740 |
| Cataracts | female | Post- | 1.3211215 | 1.7805792 | 2.3692324 |
| Hip, femoral fracture | female | Pre- | 1.1725855 | 1.1984807 | 1.2371155 |
| Hip, femoral fracture | female | Epi- | 1.0211668 | 1.0241057 | 1.0283894 |
| Hip, femoral fracture | female | Post- | 1.1548769 | 1.1779192 | 1.2122096 |

